# Post-vaccination expansion of extrafollicular Th10 and regulatory Tfr cells distinguishes strong from weak influenza vaccine responses in older adults

**DOI:** 10.64898/2026.07.02.26357118

**Authors:** Avinash S. Mahajan, Sathyabaarathi Ravichandran, Radu Marches, Yilmaz Yucehan Yazici, Sean Nelson, Teresa Aydillo-Gomez, Kshitija Kshitija, Amaya Rojo-Fernandez, Djamel Nehar-Belaid, Lisa Kenyon-Pesce, Daniel Klimes, Haebeen Jung, Peter Sage, Virginia Pascual, Patrick Wilson, Adolfo García-Sastre, Jacques Banchereau, George A. Kuchel, Duygu Ucar

**Affiliations:** The Jackson Laboratory for Genomic Medicine, 10 Discovery Drive, Farmington, CT, USA; Department of Genetics and Genome Sciences, UConn Health, Farmington, CT, USA; Gale and Ira Drukier Institute for Children’s Health and Department of Pediatrics, Weill Cornell Medicine, New York, USA; Department of Microbiology, Icahn School of Medicine at Mount Sinai, New York, USA; Global Health and Emerging Pathogens Institute, Icahn School of Medicine at Mount Sinai, New York, USA; Department of Health Sciences, University of Burgos, Burgos, Spain; UConn Center on Aging, Farmington, CT, USA; Department of Immunology, UConn Health, Farmington, CT, USA; Department of Medicine, Transplantation Research Center, Division of Renal Medicine, Brigham and Women’s Hospital, Boston, MA, USA; Immunoledge LLC; Montclair, NJ, USA; Department of Pathology, Molecular and Cell-Based Medicine, Icahn School of Medicine at Mount Sinai, New York, USA; Department of Medicine, Division of Infectious Diseases, Icahn School of Medicine at Mount Sinai, New York, USA; The Tisch Cancer Institute, Icahn School of Medicine at Mount Sinai, New York, USA; The Icahn Genomics Institute, Icahn School of Medicine at Mount Sinai, New York, USA

## Abstract

Despite the superior efficacy of high-dose influenza vaccines, over one-third of older adults fail to respond. Yet, the mechanisms underlying this impaired vaccine responsiveness remain poorly understood. Here, we performed longitudinal profiling of older adults (n=60) receiving high-dose influenza vaccination to identify immune programs associated with vaccine responsiveness. Strong responders exhibited a primed baseline immune state characterized by elevated plasma cytokines and chemokines, followed by enhanced IFN-γ responses and coordinated transcriptional and epigenetic activation of cDC2 cells at day 1. By day 7, CD4⁺ T-cell trajectories diverged: strong responders preferentially expanded influenza-specific activated cTfh1 (*CXCR5^+^ CXCR3^+^ ICOS^+^ CD38^+^*) and influenza-specific Th10 (*CXCR5⁻ CXCR3^+^ PD1^+^ IL10⁺*) cells, whereas weak responders expanded regulatory cTfr (*CXCR5⁺ FOXP3⁺*) cells. Th10 expansion correlated with plasmablast and antibody responses and was independently validated in a larger influenza vaccination cohort, including younger adults. Functionally, Th10 cells promoted memory B-cell differentiation into plasmablasts and production of influenza-specific IgGs. TCR analyses revealed minimal clonal overlap between Th10 and cTfh1 cells. Together, these findings identify divergent helper and regulatory CD4⁺ T cell programs associated with vaccine responsiveness and establish Th10 cells as a previously unrecognized component of vaccine-induced humoral immunity.

## INTRODUCTION

Despite widespread vaccination^1^, influenza remains a major cause of hospitalization and mortality among older adults (≥65 years). To improve protection in this vulnerable population, high-dose influenza vaccines containing four-fold higher antigen content have been developed and shown to reduce influenza-related illness, hospitalization, and mortality^2,3^. However, even after high-dose vaccination^4^, over one-third of older adults fail to mount robust antibody responses, and the immune mechanisms underlying these poor responses remain unknown.

Aging is accompanied by extensive remodeling of both innate and adaptive immunity, including altered antigen presentation, chronic low-grade inflammation, and impaired T and B cell responses^5–18^.Yet aging does not uniformly impair immunity. Some older adults generate durable antibody responses following vaccination, while others fail to respond despite receiving identical vaccines, suggesting that distinct immune programs underlie successful and unsuccessful vaccine responses in aging. Systems vaccinology studies have identified baseline inflammatory states, early interferon responses, circulating Tfh and plasmablast expansion as correlates of vaccine-induced immunity^19–24^. However, most of these studies have focused on younger healthy populations receiving standard-dose vaccines. Whether these signatures extend to older adults, particularly in the context of high-dose vaccination, remains unclear.

Here, we conducted an in-depth study of older adults (n=60) receiving high-dose quadrivalent influenza vaccine (Fluzone HD) by profiling antibody titers, plasma proteomics, single-cell transcriptomics with paired TCR sequencing^25^, chromatin accessibility profiling (DOGMA-seq)^26^, and CITE-seq^27^ of circulating Tfh subsets. We identified coordinated immune trajectories that distinguish strong and weak responders, including a primed baseline immune state and enhanced interferon and cDC2 responses at day 1. Strong responders preferentially expanded influenza-specific activated *CXCR5^+^ CXCR3^+^ ICOS^+^ CD38^+^* cTfh1 and *CXCR5⁻ CXCR3^+^ PD1^+^ IL10⁺* Th10 cells, whereas weak responders expanded regulatory *CXCR5⁺ FOXP3⁺* cTfr cells at day 7. Together, these findings define divergent helper and regulatory T cell programs associated with successful and unsuccessful influenza vaccine responses in older adults.

## RESULTS

### Study design and identification of strong and weak vaccine responders

To define immune determinants of influenza vaccine responsiveness in aging, we enrolled 60 community-dwelling older adults (65-92 years) during the 2022–2023 season and administered high-dose quadrivalent inactivated influenza vaccine containing A/Victoria/2570/2019, A/Darwin/9/2021 (A/H3N2), B/Austria/1359417/2021 (B/Austria), B/Phuket/3073/2013 (B/Phuket) strains (Fluzone HD; four-fold higher antigen content than standard-dose formulations) (**Fig. 1a**). The cohort comprised 36 women and 24 men, with comparable age, body mass index (BMI), and Rockwood frailty index^28^(**Fig. 1b**).

**Figure 1.**
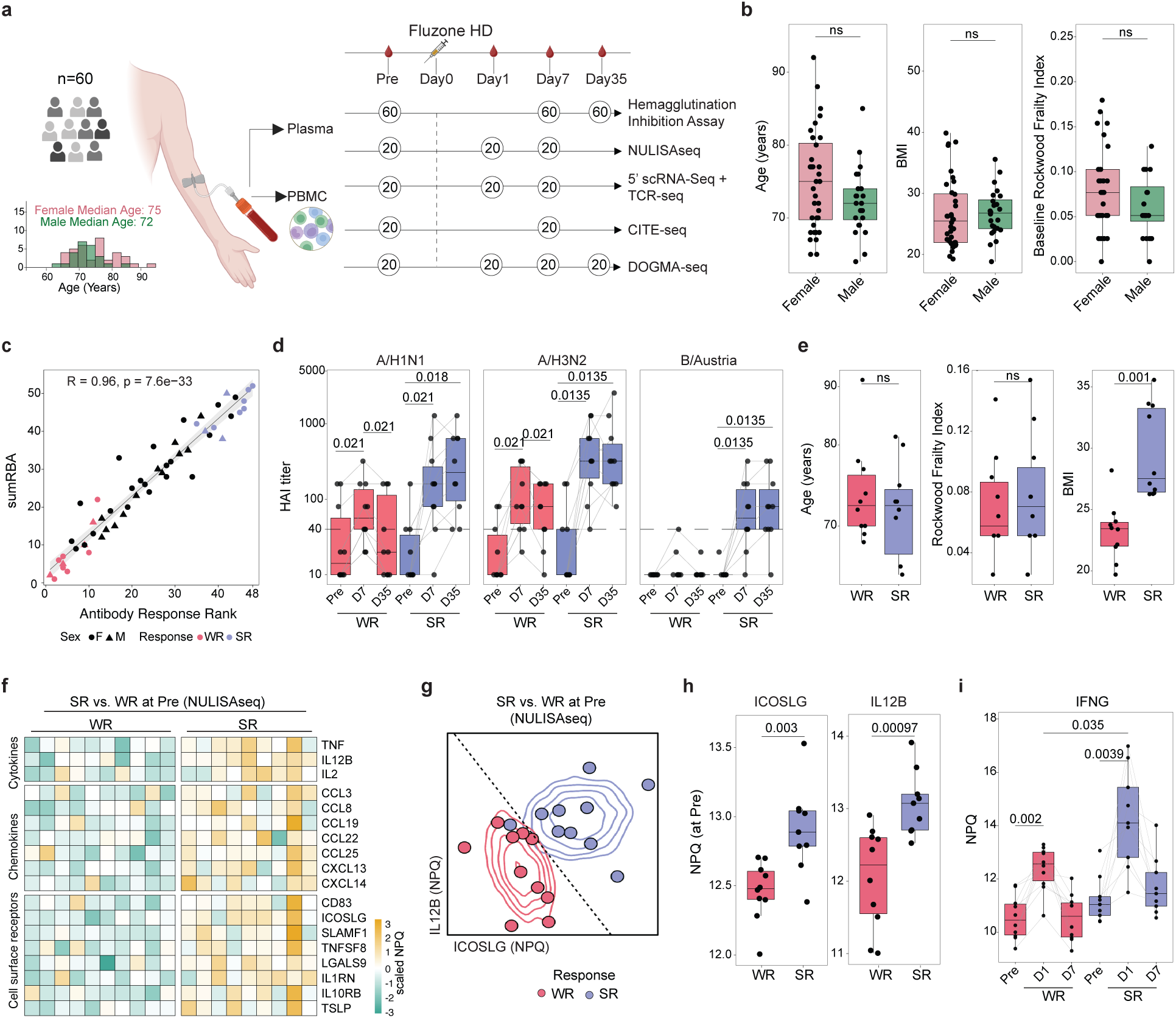
Study design and Responder stratification. a, Schematic of the study design. Sixty older adults (36 females, 24 males, age range 65–92 years) received Fluzone high-dose (HD) vaccine (2022–2023 season). Plasma collected at pre-vaccination (Pre) and days 7 and 35 were used to measure hemagglutination inhibition (HAI) titers against all four vaccine strains (n = 60). Ten strong responders (SR) and 10 weak responders (WR), selected by multivariate antibody response rank, were profiled using NULISAseq (250-plex inflammation panel). PBMCs from SR and WR (n = 10 each) were used for 5′ scRNA-seq with paired TCR-seq (Pre, days 1 and 7); CITE-seq of sorted cTfh cells (Pre, day 7); and DOGMA-seq (Pre, days 1, 7, and 35). **b,** Box plots comparing age, BMI, and Rockwood Frailty Index between females and males. **c,** Spearman correlation between antibody response rank and sumRBA (sum residual after baseline adjustment). **d,** HAI titers at Pre, days 7 and 35 in WR (n = 10) and SR (n = 10) for influenza strains A/H1N1, A/H3N2, and B/Austria. **e,** Age, Rockwood Frailty Index, and BMI compared between WR and SR. **f,** Heatmap of plasma proteins significantly elevated in SR (n = 9) versus WR (n = 10) at baseline, grouped by cytokines, chemokines, and soluble cell surface receptors. **g,** Bivariate density plot of baseline ICOSLG and IL12B NPQ values for WR (n = 10) and SR (n = 9); dashed line indicates the binomial logistic regression decision boundary. **h,** Baseline NPQ values of ICOSLG and IL12B in WR and SR. **i,** IFN-γ NPQ values at Pre, days 1 and 7 in WR and SR. Box plots show median and IQR (25th–75th percentile); whiskers extend to ±1.5 × IQR. Exact p-values are shown; ns, not significant. Wilcoxon signed-rank test (two-sided; d, i); Mann–Whitney U test (two-sided; b, e, h); Spearman correlation (c). Schematic in **a** created with BioRender.com.

Participants were free of active infection and were not receiving systemic immunosuppressive or immunomodulatory therapy at enrollment (**Table S1**). Blood samples were collected before vaccination (baseline) and on days 1, 7, 35 after vaccination. Plasma was used for hemagglutination inhibition (HAI) assays and proteomic profiling (NULISAseq^29^). Peripheral blood mononuclear cells (PBMCs) were analyzed by 5′ scRNA-seq/TCR-seq, DOGMA-seq^26^, while sorted cTfh were analyzed by CITE-seq^27^ (**Fig. 1a, Table S1**).

We quantified antibody responses to Fluzone HD before (Pre) and after vaccination (days 7, 35) using HAI assays for all four vaccine strains (**Table S1**). HAI titers increased following vaccination, with substantial inter-individual variability in both the magnitude (fold-change in HAI titers) and breadth (number of vaccine strains eliciting a response) of responses across vaccine strains (**Fig. S1a, b**). To capture donor-level heterogeneity in vaccine responses, we integrated both the magnitude and breadth of antibody responses using our established multivariate ranking framework^30^ (**Table S1**), which significantly correlated with the independently derived maximum Residual after Baseline Adjustment (maxRBA) metric^31^ (R = 0.96, p = 7.6e-33; **Fig. 1c**).

Because pre-existing antibody titers can limit the ability to achieve seroconversion, therefore we distinguished seroprotection (baseline HAI ≥40) from seroconversion (≥4-fold increase in day 35 titers relative to baseline) according to FDA criteria^32,33^. To avoid misclassifying individuals with pre-existing immunity as vaccine non-responders, donors with baseline seroprotection against two or more vaccine strains who failed to seroconvert were excluded from responder classification. Using this approach, individuals who have high responsiveness ranks and who seroconverted to two or more strains were classified as strong responders (SR), whereas those responding to one or no strains were classified as weak responders (WR). 27 out of 60 individuals (45%) failed to mount a strong response in our cohort, while 4 individuals had high-baseline titers. We selected 10 SR and 10 WR for downstream multi-omic analyses. As expected, SR exhibited robust and sustained increases in HAI titers starting at day 7 through day 35, whereas WR displayed substantially weaker responses (**Fig. 1d, S1e**). Age, sex, frailty index, and CMV serostatus were not associated with vaccine responsiveness, while BMI positively correlated with response rank (**Fig. 1e, S1c, d**).

Together, these findings demonstrate substantial heterogeneity in antibody responses to high-dose influenza vaccination among older adults.

### SRs exhibit a ‘primed’ baseline plasma proteome and enhanced IFN-γ responses

To identify circulating immune signatures associated with vaccine responsiveness, we used NULISA-seq^29^ that quantifies 250 immune-related plasma proteins at baseline, days 1, and 7 in SR (n = 10) and WR (n = 10) (**Table S2**). At baseline, SR exhibited significantly higher abundance of 25 proteins (**Fig. 1f, S2a; Table S2),** many of which positively correlated with vaccine responsiveness rank (**Fig. S2b**). These proteins spanned diverse immune pathways, including cytokines (TNF, IL2, IL12B), chemokines (CCL3, CCL25, CXCL13), and soluble surface receptors (ICOSLG, CD83), collectively suggesting that SR have a ‘primed’ immune state before vaccination. We next used binomial logistic regression models to identify proteins most strongly associated with response status. A model incorporating only two proteins, ICOSLG and IL12B, accurately discriminated SR from WR (cross-validated AUC = 1; **Fig. 1g, h**)^34,35^. We then examined post-vaccination protein dynamics and found that SR mounted significantly stronger IFN-γ responses at day 1, a result confirmed by ELISA (**Fig. 1i, S2c; Table S2**).

Together, these findings indicate that effective vaccine responses in older adults are associated with a distinct baseline cytokine milieu that is followed by enhanced early IFN-γ production after vaccination.

### Early cDC2 activation distinguishes successful vaccine responses

To characterize early cellular responses to vaccination, we profiled PBMCs from SR and WR using 5′ scRNA-seq with paired TCR sequencing across baseline, days 1, and 7 (956,539 cells; **Fig. S3a, b; Table S3**). Unsupervised clustering followed by manual annotation with canonical markers identified eight major subsets: dendritic cells (DCs), CD14^+^ monocytes, CD16^+^ monocytes, Natural Killer (NK) cells, CD4^+^ and CD8^+^ T cells, B cells and plasmablasts. Within DCs, we identified five subsets: conventional type 1 DCs (cDC1; *CLEC9A^+^*; n=409), conventional type 2 DCs (cDC2; *CLEC10A^+^, CD14^-^*; n=3831), monocyte-derived DCs (monoDCs; *CLEC10A^+^, CD14^+^*; n=2848), plasmacytoid DCs (*IRF7^+^*; n=2905) and AXL-DCs (*AXL^+^, SIGLEC6*^+^; n=228) (**Fig. 2a**), consistent with established transcriptional definitions^36,37^.

**Figure 2.**
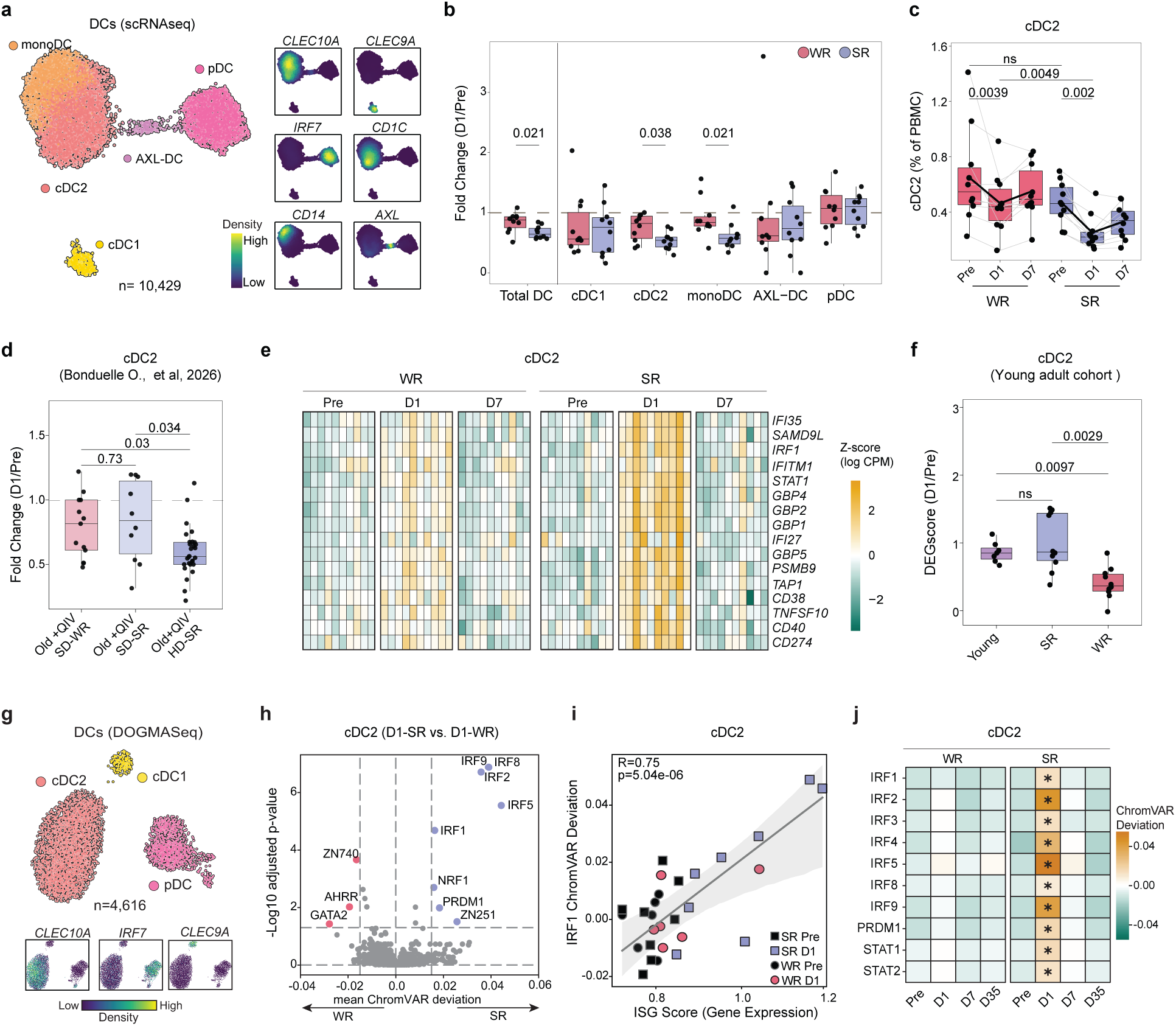
Day 1 cDC2 egress and interferon-driven transcriptional response define SR. a, UMAP of DC subsets from scRNA-seq (n = 10,429 cells) identifying five transcriptionally distinct subsets: cDC1, cDC2, monoDC, AXL-DC, and pDC. Feature density plots show expression of canonical marker genes. **b,** Box plots comparing fold change (day 1/Pre) for each DC subset between WR (n=10) and SR (n = 10). **c,** Longitudinal cDC2 frequencies (% of PBMCs) in WR (n=10) and SR (n = 10) at Pre, days 1 and 7. **d,** cDC2 fold change (day 1/Pre) in older adults from an independent cohort receiving standard-dose quadrivalent inactivated influenza vaccine classified as SR (QIV SD-SR) or WR (QIV SD-WR), or high-dose QIV classified as SR (QIV HD-SR), quantified from flow cytometry. **e,** Heatmap of Z-score normalized expression of genes differentially expressed (day 1 versus Pre) in cDC2s of SR. Full list of differentially expressed genes is provided in **Table S3**. **f,** Box plots comparing the transcriptional DEG score (day 1/Pre) in cDC2s across young adults receiving Fluzone standard dose (n = 7), SR (n = 10), and WR (n = 10). **g,** UMAP of DOGMA-seq data (n = 4,616 cells) with cDC1, cDC2, and pDC subsets; feature plots show selected marker genes. **h,** Volcano plot of ChromVAR deviations in cDC2s comparing SR versus WR at day 1; IRF family motifs are significantly enriched in SR. **i,** Spearman correlation between transcriptome-based ISG score and IRF1 ChromVAR deviation score in cDC2s. **j,** Heatmap of ChromVAR deviation scores for IRF and STAT family transcription factor motifs in cDC2s from WR and SR across Pre, days 1, 7, and 35; asterisks indicate significant enrichment in SR at day 1. Box plots display the median and IQR (25th–75th percentile); whiskers extend to ±1.5 × IQR. Solid lines in boxplot indicate the group mean at each timepoint. Exact p-values are shown; ns, not significant. Wilcoxon signed-rank test (two-sided; c); Mann–Whitney U test (two-sided; b, c, d, f); Spearman correlation (i).

No differences in DC frequencies were observed at baseline. However, by day 1, total circulating DC frequencies declined significantly in SR relative to WR, driven primarily by reductions in cDC2 (p = 0.038) and monoDC (p = 0.021) populations (**Fig. 2b, c, S3c**). The magnitude of decline in these subsets negatively correlated with vaccine responsiveness rank (**Fig. S3d**). We further analyzed flow cytometry data from an independent cohort of older adults receiving either standard(n=23) or high-dose(n=28) influenza vaccination during the 2021–2022 season^6^. Consistent with our cohort, day 1 reductions in cDC2 frequencies were observed specifically in SR receiving high-dose vaccine and correlated with antibody responses (**Fig. 2d, S3e**).

### cDC2s from SR mount enhanced interferon responses

We next examined transcriptional responses within DC subsets. In SR, differential expression analysis comparing day 1 to baseline samples revealed robust transcriptional activation in cDC2s (79 up, 13 down) and monoDCs (91 up, 18 down) (FDR 5%). In contrast, in WR, there were no significant transcriptional changes across any DC subsets (**Fig. S3f, Table S3**). Upregulated genes in cDC2s and monoDCs included canonical interferon stimulated genes (ISGs: *IFI35, IRF1, IFITM1, STAT1, GBP5, IFI27*) and antigen processing and presentation genes (*TAP1, PSMB9, CD40, CD274/PD-L1*) (**Fig. 2e, S3g, Table S3**). Confirming these observations, gene-set enrichment analysis comparing SR and WR at day 1 in cDC2s demonstrated significant enrichment of IFN-γ response, IFN-α response, TNFα signaling via NF-κB, and antigen processing and presentation pathways (**Fig. S3h, Table S3**). These transcriptional responses were transient and largely resolved by day 7.

To compare these transcriptional responses to that of younger adults, we analyzed an independent cohort comprising younger adults who received Fluzone SD (n=7, age range = 22-39 years). Younger adults mounted a strong ISG response at day 1 in cDC2s, which was comparable in magnitude to the responses of SR in the high-dose vaccine cohort. In contrast, older WR exhibited substantially attenuated transcriptional responses compared to SR and younger adults receiving standard dose (**Fig. 2f**). This indicates that successful high-dose vaccination in older adults induce transcriptional responses comparable to younger adults receiving standard-dose vaccination.

To investigate the epigenetic basis of these transcriptional responses, we analyzed longitudinal DOGMA-seq data. ChromVAR^38^ analysis revealed increased accessibility of interferon regulatory factor (IRF) motifs in cDC2s from SR at day 1, consistent with their enhanced transcriptional interferon responses (**Fig. 2g,h, S3i**). Notably, IRF1, STAT1, STAT2, and IRF9 exhibited coordinated increases in both gene expression and motif accessibility at day 1 in cDC2 (**Fig. 2i, j**). Similarly, SR mounted more significant ISG responses in monocytes compared to WR both epigenetically and transcriptionally (**Fig. S4a-j, Table S4**).

In summary, successful vaccine responses are associated with a rapid IFN-γ–driven activation program in cDC2s, that resembles responses observed in younger adults.

### SRs expand IgG1⁺ plasmablasts

We analyzed 61,871 B cells and identified naïve (*IGHD*⁺; n = 43,334), memory (*CD27*⁺; n = 16,195), and plasmablast (*JCHAIN*⁺, *MZB1*⁺, *PRDM1*⁺; n = 2,342) populations (**Fig. 3a**). While B cell subset frequencies were comparable between SR and WR at baseline, plasmablast frequencies increased at day 7, only in SR^39^ (p = 0.03; **Fig. S5a**).

**Figure 3.**
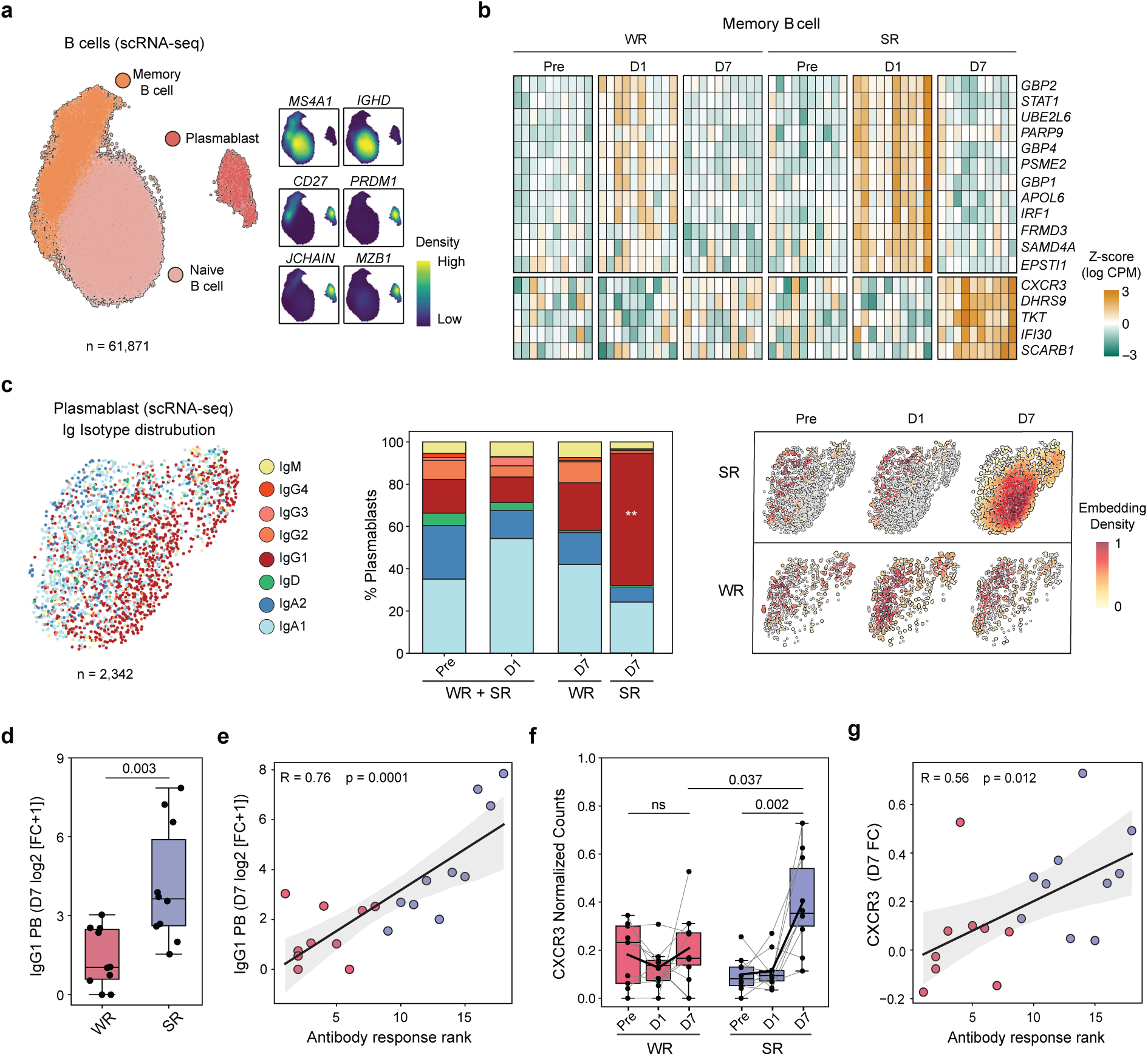
B cell activation and IgG1 plasmablast expansion in SR upon vaccination. a, UMAP of B-lineage cells from scRNA-seq (n = 61,871 cells) identifying three transcriptionally distinct subsets: naïve B cells, memory B cells, and plasmablasts. Feature density plots show expression of selected marker genes. **b,** Heatmap of Z-score normalized expression of the top 30 genes differentially expressed (days 1 and 7 versus Pre) in memory B cells. Full list of differentially expressed genes is provided in **Table S5**. **c,** Left: UMAP of plasmablasts (n = 2,342 cells) identifying isotype-defined clusters. Center: Stacked bar plots showing the distribution of plasmablast isotype clusters across Pre, days 1 and 7 in older adults (n = 20). Right: embedding density of timepoints overlaid on plasmablast UMAP split by response group across timepoints. **d,** Box plots comparing fold change (day 7/Pre) of IgG1 plasmablasts between WR (n=10) and SR (n = 10). **e,** Spearman correlation between day 7 IgG1 plasmablast fold change and antibody response rank. **f,** Longitudinal CXCR3 expression in plasmablasts from WR(n = 9) and SR (n = 10) at Pre, days 1 and 7; solid lines indicate the group mean at each timepoint. **g,** Spearman correlation between day 7-fold change in CXCR3 expression in plasmablasts and antibody response rank. Box plots display the median and IQR (25th–75th percentile); whiskers extend to ±1.5 × IQR. Solid lines in boxplot indicate the group mean at each timepoint. Exact p-values are shown; ns, not significant. Wilcoxon signed-rank test (two-sided; f); Mann–Whitney U test (two-sided; d, f); Spearman correlation (e, g).

Differential expression analysis revealed strong ISG responses in SR at day 1, as observed in myeloid cells (**Fig. S5b, c**). In addition, at day 7, memory B cells from SR upregulated *CXCR3* (**Fig. 3b**), consistent with IFN-γ–induced CXCR3 upregulation on memory B cell^40,41^. Further subclustering identified DN2 cells (*CD27⁻, IgD⁻, FCRL5⁺, TBX21⁺, ITGAX⁺*) and an activated memory B-cell population characterized by expression of *CD27, CR2, ITGAX*, *TBX21*, and *FCRL5*^42^ (**Fig. S5d**). DN2 cells did not expand upon vaccination, whereas the activated memory B cell subset expanded in both SR and WR (**Fig. S5e, f**).

To characterize the isotype composition of the plasmablast response, we examined immunoglobulin isotype expression at the single-cell level. Upon vaccination, only IgG1⁺ plasmablasts significantly expanded at day 7 selectively in SR (p = 0.003; **Fig. 3c, d**). This IgG1⁺ plasmablast expansion strongly correlated with vaccine responsiveness rank (R = 0.76, p = 0.0001; **Fig. 3e**). In SR, CXCR3 was upregulated in plasmablasts (p = 0.002) and positively correlated with antibody responses (R = 0.56, p = 0.012; **Fig. 3f, g**), a program shown to be driven by extrafollicular T helper populations during viral infection^43^.

Together, these results demonstrate that successful vaccination selectively induces IgG1⁺ plasmablast expansion in SR, accompanied by CXCR3 upregulation on both memory B cells and plasmablasts, consistent with an IFN-γ–driven activation program.

### Influenza-specific cTfh1 expand in SR

CD4⁺ and CD8⁺ T cells from SR also exhibited strong transcriptional ISG responses at day 1 that were largely absent in WR (**Fig. S6, Table S5**). Reclustering of memory CD4⁺ T cells identified multiple helper populations: T helper (Th)1-like (*CXCR3*^+^, *IFNG-AS1*^+^, *GZMK*^+^), Th2 (*GATA3*^+^, *GATA3-AS1*^+^, *PTGDR2*^+^), Th17 (*CCR6*^+^, *CTSH*^+^, *RORC*^+^), Th22 (*CCR10*^+^), central memory (*SELL*^+^), CD38^+^ central memory, T follicular helper cells (Tfh: *CXCR5*^+^), TEMRA (*GZMB*^+^), and IL10^+^ *(IL10*^+^, *PD1*^+^) cluster (**Fig. S7a**). All CD4⁺ memory subsets, except TEMRAs declined significantly in frequencies at day 1 in SR (**Fig. S7b**). cTfh cells are established correlates of influenza vaccine responses, with activated ICOS⁺CD38⁺ cTfh cells showing the strongest associations with antibody magnitude^44,45^.

We further resolved the cTfh compartment and identified an activated *ICOS⁺ CD38⁺ CXCR3⁺* population, referred to hereafter as activated cTfh1 cells (**Fig. 4a**). Frequencies of activated cTfh1 cells increased significantly at day 7 in SR but not WR (p = 0.02; **Fig. 4b**). The magnitude of their expansion positively correlated with vaccine responsiveness rank (R = 0.52, p = 0.019; **Fig. S7c**) and IgG1⁺ plasmablast expansion (R = 0.51, p = 0.022; **Fig. 4c**). Flow cytometry analysis confirmed preferential expansion of activated cTfh1 cells in SR (p = 0.037; **Fig. 4d, Table S6**).

**Figure 4.**
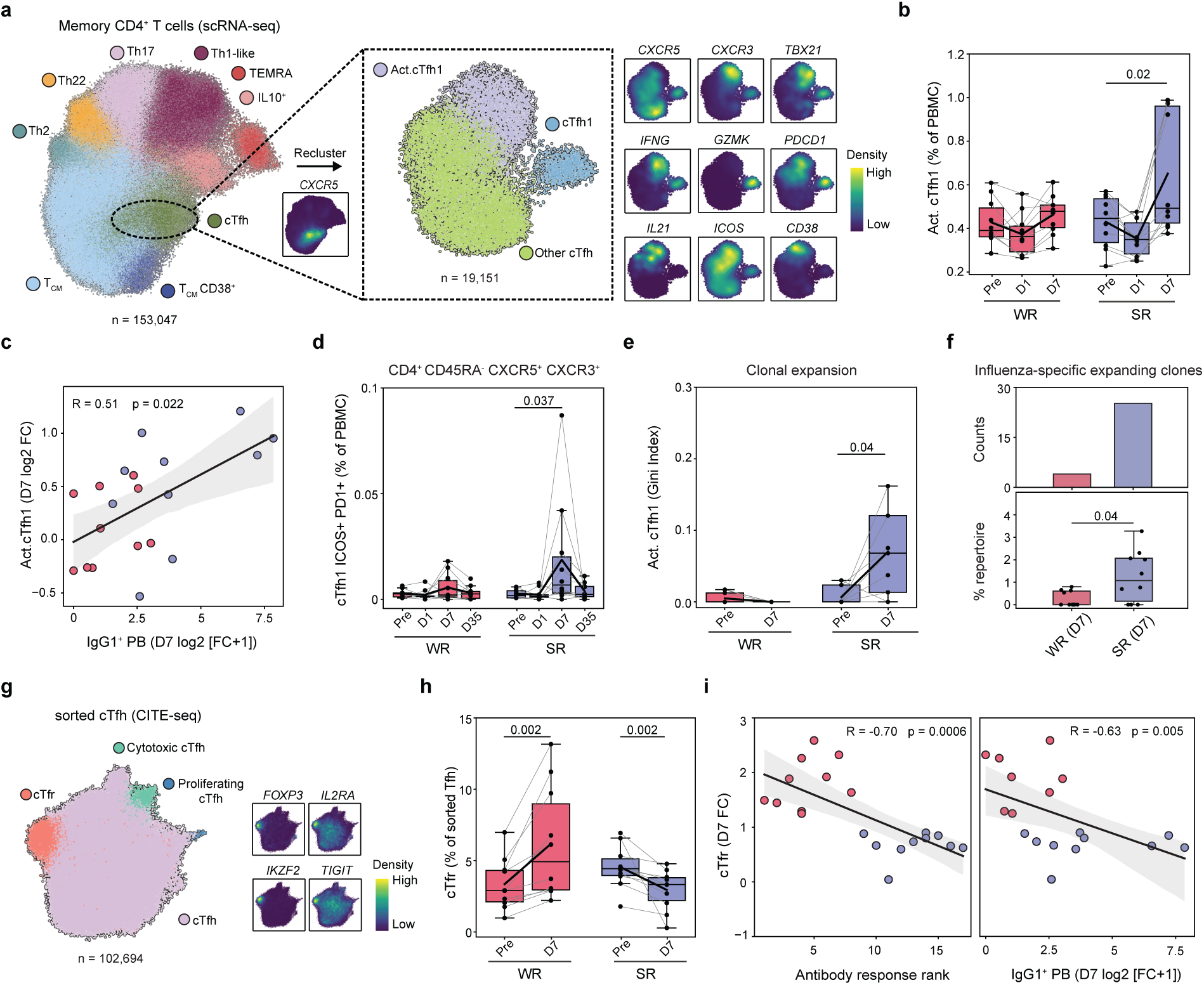
Expansion of Activated cTfh1 in SR, and cTfr in WR at day 7. a, Left: UMAP of memory CD4⁺ T cells from scRNA-seq (n = 153,047 cells) identifying transcriptionally distinct subsets. Right: UMAP of reclustered cTfh cells (n = 19,151 cells) identifying cTfh subsets. Feature density plots show expression of selected cTfh1 and activation-associated marker genes. **b,** Longitudinal activated cTfh1 frequencies in WR (n=10) and SR (n = 10) at Pre, days 1 and 7. **c,** Spearman correlation between day 7 activated cTfh1 expansion and day 7 IgG1 plasmablast expansion. **d,** Longitudinal CD4⁺CD45RA⁻CXCR5⁺CXCR3⁺PD-1⁺ cell frequencies quantified by flow cytometry in WR (n=10) and SR (n = 10) at Pre, days 1 and 7. **e,** Activated cTfh1 clonal expansion, measured by Gini index, at Pre and day 7 in WR and SR (n = 10 each). **f,** Top: bar plot showing the number of influenza-specific expanding activated cTfh1 clones at day 7 in WR and SR. Bottom: percentage of day 7 influenza-specific expanding activated cTfh1 clones in WR(n=10) and SR (n = 10). **g,** UMAP of sorted cTfh cells from CITE-seq (n = 102,694 cells); feature density plots show selected cTfr marker genes. **h,** Longitudinal cTfr cell proportions in WR (n=10) and SR (n = 10) at Pre, days 1 and 7. **i,** Spearman correlations between day 7 cTfr expansion and antibody response rank (n = 20) or day 7 IgG1 plasmablast expansion (n = 18) from scRNA-seq. Solid lines indicate the group mean at each timepoint. Box plots display the median and IQR (25th–75th percentile); whiskers extend to ±1.5 × IQR. Solid lines in boxplot indicate the group mean at each timepoint. Exact p-values are shown; ns, not significant. Wilcoxon signed-rank test (two-sided; b, d, e, h); Mann–Whitney U test (two-sided; b, d, e, f, h); Spearman correlation (c, i).

We analyzed paired TCR-seq data (381,396 T cells in total; mean of 6,356 cells per sample). At day 7, activated cTfh1 cells displayed clonal expansion in SR (p = 0.04) but not in WR (**Fig. 4e**). To determine whether the expanding cTfh1 cells were influenza-specific, we curated 61,438 putative–influenza-specific TCR sequences from public databases including experimentally validated sequences^46–51^ (**Table S7**). SRs harbored 25 expanding influenza-specific activated cTfh1 clonotypes at day 7, compared to 3 in WR. The frequency of these clones at the donor level was significantly higher in SR (p = 0.04), despite no differences at baseline (**Fig. 4f**).

Together, these findings identify clonal expansion of influenza-specific activated cTfh1 cells as a hallmark of successful responses to high-dose influenza vaccination in older adults, extending prior observations in younger adults.

### cTfr cells expand in WR and contract in SR

To gain higher resolution within the cTfh compartment, we sorted CD4^+^ CXCR5^+^ CD45RA^-^ cells from 20 donors at baseline and day 7 (10 SR and 10 WR) and performed CITE-seq using surface markers (CCR6, CD38, CXCR3, HLA-DR, ICOS, PD1) (**Fig. S8a**). In alignment with 5’ scRNAseq data, SR displayed elevated CD38 surface protein levels at day 7 in cTfh1, whereas WR showed no significant changes (**Fig. S8c, d**).

This increased resolution also revealed a T follicular regulatory (cTfr) population expressing *FOXP3*, *IL2RA*, *IKZF2*, and *TIGIT* (**Fig. 4g, S8b**)^52^. Remarkably, cTfr frequencies increased in WR upon vaccination (p = 0.002; **Fig. 4h**). In contrast, their frequencies declined in SR following vaccination (p = 0.002; **Fig. 4h**). The magnitude of cTfr expansion negatively correlated with vaccine responsiveness rank (R = −0.70, p = 0.0006) and IgG1⁺ plasmablast expansion (R = −0.63, p = 0.005) (**Fig. 4i**), suggesting that cTfr expansion in WR may actively limit the humoral response following vaccination^44,52^.

Together, these findings reveal a striking divergence in the cTfr response following vaccination, cTfr expansion in WR inversely associated with antibody output.

### Influenza-specific Th10 cells expand in SR and promote plasmablast differentiation

Within the memory CD4⁺ T cell compartment, we identified a distinct IL10^+^ population and further resolved it into Th10 (*CXCR3*⁺ *CXCR5*^-^ *IL10*⁺ *CD38*⁺ *PD1*⁺), cTfh10 (*CXCR5*⁺ *CXCR3*⁺ *IL10*⁺ *PD1*⁺), and T_EM_ HLA-DR^+^ effector memory populations (**Fig. 5a**). Th10 cells are a *CXCR5*^-^ *CXCR3*⁺ *IL10*⁺ *PD1^+^* helper T cell population, recently described in autoimmune contexts that promote plasmablast differentiation and antibody production through extrafollicular pathways^53,54^. Their contribution to vaccine-induced immunity has not been examined.

**Figure 5.**
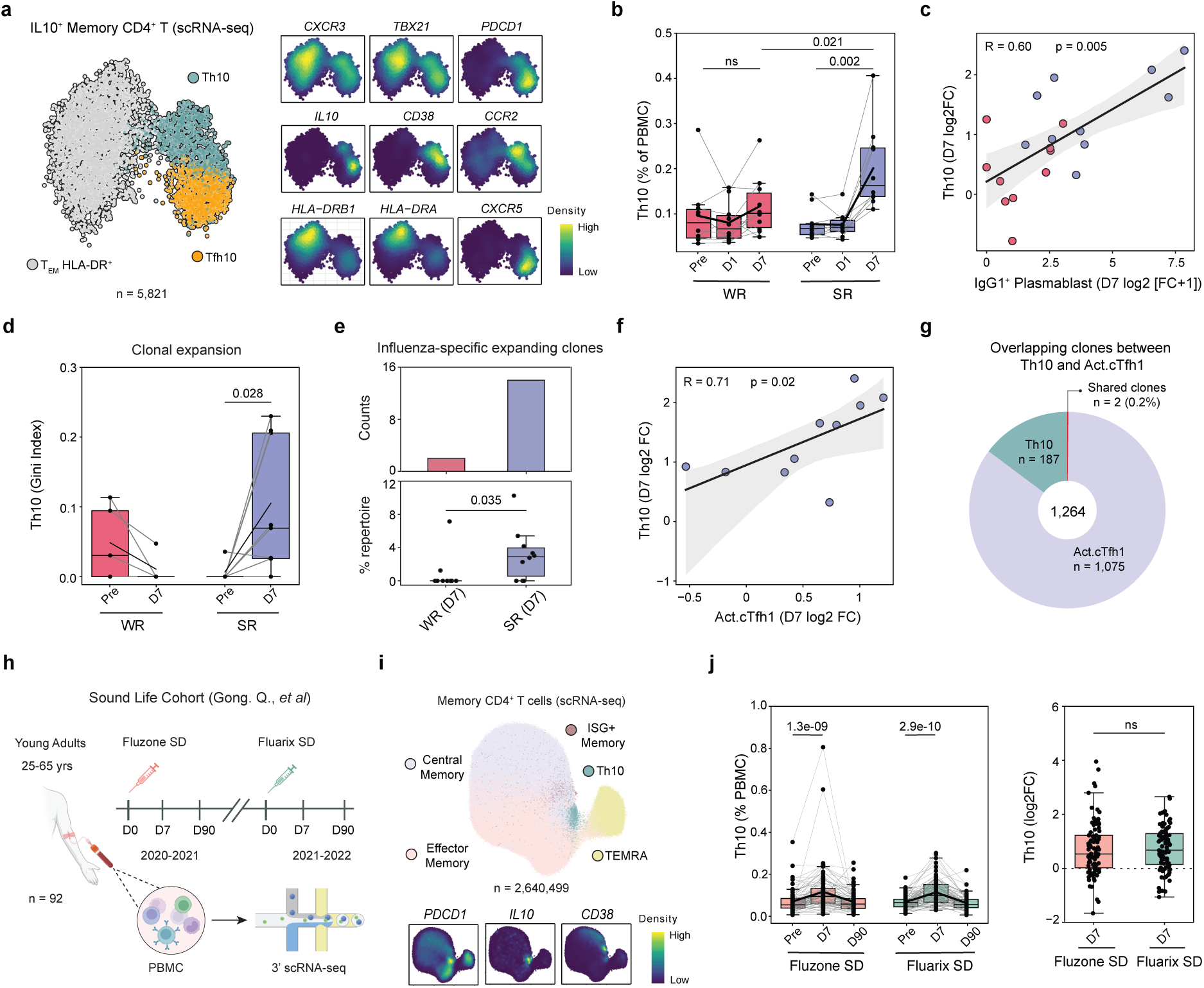
Th10 expansion is linked to plasmablast expansion and strong antibody responses. a, UMAP of memory CD4⁺ T cell subclusters from scRNA-seq (n = 5,821 cells), including Th10, Tfh10, and HLA-DR⁺ cells. Feature density plots show expression of selected marker genes. **b,** Longitudinal Th10 cell frequencies in WR and SR (n = 10 each) at Pre, days 1 and 7. **c,** Spearman correlation between day 7 Th10 expansion and day 7 IgG1 plasmablast expansion. **d,** Th10 clonal expansion, measured by Gini index, at Pre and day 7 in WR (n=10) and SR (n = 10). **e,** Top: bar plots showing the number of influenza-specific expanding Th10 clones at day 7 in WR and SR. Bottom: percentage of day 7 influenza-specific expanding Th10 clones in WR (n=10) and SR (n = 10). **f,** Spearman correlation between day 7 activated cTfh1 expansion and day 7 Th10 expansion in SR. **g,** Donut plot showing the proportion of unique and shared clonotypes between Th10 and activated cTfh1 cells, quantified by clone number. **h,** Overview of the independent Sound Life cohort used for validation, including 92 healthy adults receiving standard-dose influenza vaccines across the 2020–2021 and 2021–2022 seasons. **i,** UMAP of memory CD4⁺ T cell subsets from the Sound Life younger adult cohort (n = 2,640,499 cells); feature density plots show expression of selected marker genes. **j,** Longitudinal Th10 frequencies and day 7 Th10 fold change after standard-dose influenza vaccination across two consecutive seasons. Box plots display the median and IQR (25th–75th percentile); whiskers extend to ±1.5 × IQR. Solid lines indicate the group mean at each timepoint. Exact p-values are shown; ns, not significant. Wilcoxon signed-rank test (two-sided; b, d, j); Mann–Whitney U test (two-sided; b, d, e, j); Spearman correlation (c, f).

Th10 frequencies increased significantly at day 7 in SR but not WR (p = 0.002; **Fig. 5b**). Accordingly, SR exhibited higher Th10 frequencies than WR at day 7 (p = 0.021; **Fig. 5b**). The magnitude of Th10 expansion positively correlated with antibody response rank (R = 0.66, p = 0.002) and IgG1⁺ plasmablast expansion (R = 0.60, p = 0.005) (**Fig. 5c, S7d**). Flow cytometry analysis confirmed expansion of CD4^+^ CXCR5^-^ CXCR3^+^ PD1^+^ (Th10) cells at day 7 in SR (**Fig. S7e, Table S6**). We next analyzed paired TCR-seq data. Th10 cells showed significant clonal expansion at day 7 in SR but not WR (p = 0.028; **Fig. 5d**).

To determine whether the expanding Th10 cells were influenza-specific, we compared them against putative–influenza-specific TCR sequences (**Table S7**). Baseline frequencies of influenza-specific Th10 clonotypes were comparable between groups, but SR exhibited 14 expanding influenza-specific clonotypes at day 7, substantially higher than WR. Moreover, the frequency of these expanding clones was higher in SR (p = 0.035; **Fig. 5e**).

Among all memory CD4⁺ T cell subsets profiled, activated cTfh1 and Th10 were the only populations that expanded significantly at day 7 in SR. Expansion of Th10 and activated cTfh1 cell frequencies positively correlated in SR (R = 0.71, p = 0.02; **Fig. 5f**), but not in WR (R = −0.3, p = 0.4; **Fig. S7f**). We next examined whether they share clones. Overall, there were a total of 1,264 clones in Th10 and cTfh1 cells. Only 2 clones were shared by the two populations (0.2%; **Fig. 5g**), suggesting that cTfh1 and Th10 represent distinct antigen-driven helper T cell populations expanding together in SR.

To assess whether Th10 expansion is a general feature of influenza vaccine responses, we analyzed an independent cohort of young (25-35 years) and middle-aged adults (55-65 years) vaccinated across two influenza seasons with Fluzone SD (n = 92) and Fluarix SD (n = 84)^55^. Single-cell analysis of the CD4^+^ memory T cell (n = 2,640,499) compartment revealed Th10 cells (n = 11,071 cells) based on previously described markers^53,54^ (**Fig. 5h, i, S9a**). Th10 frequencies increased significantly at day 7 following vaccination for both age groups. In addition, this expansion was observed in both vaccine formulations (Fluzone SD: p = 1.3e-09, Fluarix SD: p = 2.9e-10; **Fig. 5j, S9b**). In the B cell compartment, plasmablast proportions increased at day 7 (Fluzone SD: p = 2.4e-06, Fluarix SD: p = 0.001) (**Fig. S9c-e**) and positively correlated with Th10 expansion (Fluzone SD: R = 0.35, p = 0.0006; Fluarix SD: R = 0.33, p = 0.002) (**Fig. S9f, g**).

To determine whether vaccine-induced Th10 cells directly support humoral responses, we performed B-cell co-culture experiments using sorted cTfh (CD4^+^ CD45RA^-^ CXCR5^+^) and Th10 (CD4^+^ CD45RA^-^ CXCR5^-^ CXCR3^+^ PD1^+^) and autologous memory B cells (CD19^+^ CD27^+^ IGHD^-^) from PBMCs of six donors at day 7 post influenza vaccination (**Fig. 6a, S10**). Both Th10 and cTfh cells promoted robust and comparable differentiation of memory B cells into plasmablasts (**Fig. 6b, c, Table S8**). ELISA measurements of culture supernatants showed that Th10 and cTfh cells also induced comparable levels of Fluzone-reactive IgG (**Fig. 6d, Table S8**), demonstrating that Th10 cells provide B-cell help of a magnitude similar to canonical cTfh cells.

**Figure 6.**
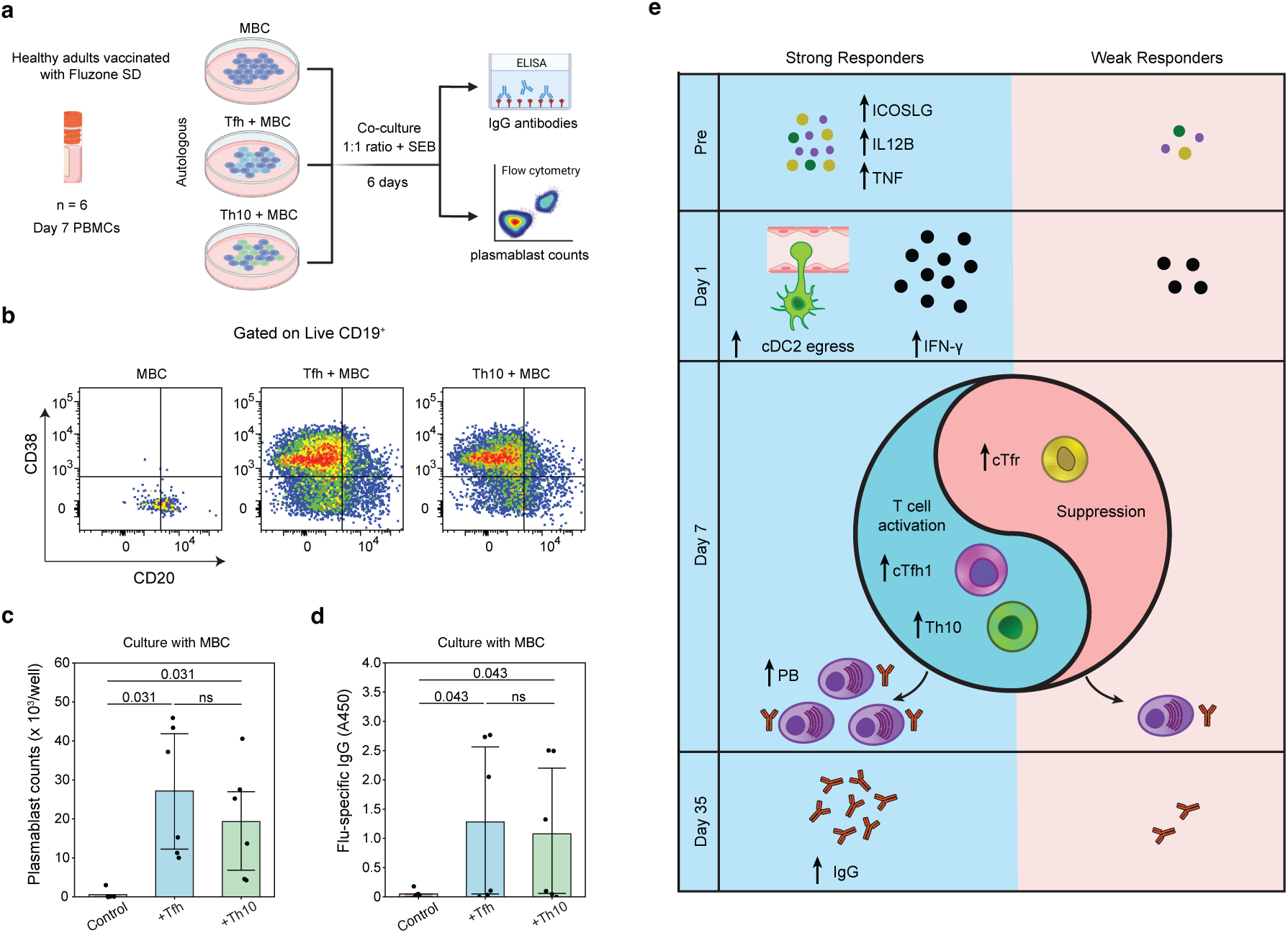
Th10 promotes plasmablast differentiation and influenza-specific IgG production. a, Schematic overview of autologous coculture experiments using memory B cells, Th10 cells, and cTfh cells sorted from day 7 post-vaccination PBMCs from healthy adults (n = 6; see Methods). Memory B cells were cultured alone or cocultured with the indicated CD4⁺ T helper cell subsets at a 1:1 ratio (20,000 cells per celltype) in the presence of Staphylococcus enterotoxin B for 6 days. **b,** Representative flow cytometry plots showing CD38 and CD20 expression in memory B cells cultured alone or with the indicated CD4⁺ T helper cell subsets, used to identify plasmablasts. **c,** Bar plots showing plasmablast counts after 6 days of culture across the indicated conditions in healthy adults (n = 6). Note that both cTfh and Th10 cocultures induced robust plasmablast differentiation. **d,** Bar plots showing influenza-specific IgG antibody levels quantified from coculture supernatants across the indicated conditions in healthy adults (n = 6). Note that cTfh and Th10 cocultures significantly increased influenza-specific IgG production relative to memory B cells cultured alone. **e,** Summary figure outlining the myeloid and adaptive signatures of influenza vaccine responses in SR and WR. Bar plots display the median and IQR (25th–75th percentile); whiskers extend to ±1.5 × IQR. Exact p-values are shown; ns, not significant. Wilcoxon signed-rank test (two-sided; c, d).

Together, these findings reveal a previously unappreciated role for Th10 cells in vaccine-induced immunity, demonstrating their capacity to support plasmablast differentiation and influenza-specific IgG production operating in parallel with canonical cTfh cells.

## DISCUSSION

Why some older adults respond robustly to influenza vaccination while others do not, despite comparable age, health, and vaccine exposure has been a longstanding question in aging and vaccinology. Using an integrative systems immunology approach, we identify cellular and molecular determinants that distinguish strong from weak humoral responses to high-dose influenza vaccination in older adults. Our findings reveal that immune resilience in aging is not uniformly lost but is preserved in a subset of older adults through a coordinated program spanning innate activation, antigen-specific T helper expansion, and regulated antibody output, that is likely suppressed in weak responders (**Fig. 6e**).

The cellular differences between SR and WR were detected in Antigen Presenting Cells (APC) as early as day 1 post-immunization. SRs exhibited a robust transcriptional and epigenetic induction of interferon-response programs alongside rapid decline in cDC2 frequencies, suggesting egress from blood to draining lymphoid tissues. cDC2s are specialized cells that efficiently prime CD4⁺ T cell responses and support Tfh differentiation^56^. Expansion of cTfh at day 7 has been linked to vaccine-induced plasmablast responses in both younger and older adults^11,57,58^. However, heterogeneity of these responses in older adults have not been explored before. Our data establish that expansion of cTfh1 cells is a critical determinant of successful humoral responses in this population.

A central and unexpected finding of this study is the identification of vaccine-induced Th10 cells as a novel helper population in humans. Th10 cells, originally described in autoimmune settings, promote plasmablast differentiation and antibody production through IL-10-dependent mechanisms^53,54^. Their role in vaccine-induced immunity had remained unknown. Here, we demonstrate that successful vaccination is associated with expansion of influenza specific Th10 cells in responding older adults. Functional co-culture experiments established that these cells are *bona fide* B cell helpers, capable of promoting memory B cell differentiation into plasmablasts and driving secretion of influenza-specific IgG at levels comparable to canonical cTfh cells. Th10 expansion was not restricted to older adults, as we show their expansion in young and middle-aged adults vaccinated with inactivated influenza vaccines^55^. Notably, Th10 and cTfh1 expansion were correlated at the population level in SR, yet the two populations exhibited minimal TCR clonotype overlap. Another CD4^+^ population with extrafollicular function is T peripheral helper (Tph) cells (CXCR5⁻PD1^hi^) which provide B cell help through IL-21 and CXCL13 and are typically expanded in autoimmune disease and dengue infection^59,60^. Consistent with the non-inflamed context of vaccination, a distinct Tph cluster was not identified in this healthy cohort.

Unlike cTfh1 cells, Th10 cells are poised for extrafollicular rather than germinal center niches. Extrafollicular T helper-driven responses are characteristically rapid, generating plasmablasts and class-switched antibodies within days of antigen encounter, preceding the slower GC reaction by days, and can produce neutralizing antibodies of comparable protective capacity^61,62^. This kinetic advantage may be particularly relevant in the context of aging, where GC magnitude and quality are known to decline due to impaired Tfh differentiation and spatial disorganization of the GC microenvironment^9,10^. Engagement of an extrafollicular Th10-driven pathway may partially compensate for weakened GC-dependent responses, providing a rapid, memory B cell source of antibody output that is preserved in SR but absent in WR. Together, these data suggest that successful vaccination in older adults engages both germinal center (cTfh1) and extrafollicular (Th10) T helper pathways that potentially serve as a complementary humoral arm in older adults.

Another key finding of our study is the expansion of cTfr in WR and their decline in SR. cTfr expansion inversely correlated with plasmablast induction and antibody responses. Previous studies in humans show that when co-cultured with B and Tfh cells, cTfr cells constraint plasmablast expansion and IgG secretion in vitro^52^. A recent study demonstrated that cTfr cells undergo transient contraction following influenza vaccination in younger adults. Our findings extend this observation by showing that SR specifically recapitulate this young-like cTfr contraction, whereas WR exhibit the opposite, suggesting that the balance between helper and regulatory T follicular subsets is a key determinant of vaccine responsiveness in aging. The mechanisms by which cTfr expansion suppresses helper T cell function and humoral output in WR, and whether targeting this regulatory axis could enhance vaccine responses in non-responders, remain important questions for future investigation.

These findings identify multiple cellular checkpoints that distinguish strong from weak responses to high-dose influenza vaccination in aging. The baseline plasma proteomic differences between SR and WR, enriched for co-stimulatory ligands and chemokines, suggest that pre-vaccination immune priming may be a modifiable target for adjuvant-based strategies aimed at improving vaccine responsiveness^19,63–66^. The observation that SR among older adults recapitulate key features of young adult vaccine responses raises the question of what maintains immune resilience in a subset of older individuals, and whether this resilience is heritable, environmentally conditioned, or modifiable. Our study identified multiple cellular checkpoints that could be targeted to improve vaccine responsiveness in older adults.

Several limitations should be considered. First, deep multi-omic profiling was performed in a subset of participants (10 strong and 10 weak responders), and larger cohorts will be required to validate the identified immune signatures and evaluate their predictive value across diverse populations. Second, all analyses were conducted using peripheral blood.

cDC2 trafficking, lymph node priming, germinal center activity, and local T–B cell interactions could not be directly assessed and were inferred from circulating immune populations.

## Methods

### Participant recruitment

This study was conducted following approval by the UConn Health Center Institutional Review Board (22-179J-1) and registration on ClinicalTrials.gov (NCT05518500). The complete information on clinical trial registration, study protocol, data collection and outcomes can be accessed via https://clinicaltrials.gov/study/NCT05518500. All participants provided informed consent and were compensated for their time and study visits. Blood samples were obtained from 63 healthy volunteers residing in the Greater Hartford, CT, region recruited by the UConn Center on Aging.

Recruitment for the UConn Center on Aging (COA) site was conducted using two primary sources: the COA Recruitment Volunteer Registry database and the JAX COVID-19 Biorepository registry, both of which included individuals who had previously consented to be contacted about future research. IRB-approved recruitment letters or emails describing the study and providing contact information were sent to potentially eligible participants, and interested individuals contacted the COA study team directly. Additional recruitment strategies included flyers in community locations and clinics, internal UConn Health advertising (e.g., TV monitors and broadcast emails), and all materials and methods were approved by the IRB prior to use.

Inclusion criteria included participants aged 65 or older and willing to receive FDA-approved seasonal influenza vaccination. Participants were excluded if they had received any vaccine (e.g., shingles, pneumococcal, or COVID-19) within two weeks of the anticipated influenza vaccination, had already received the current season’s influenza vaccine, or had a history of severe reaction to the flu vaccine or allergy, or Guillain-Barré syndrome. Additional exclusions included significant comorbid conditions such as recent or chronic infections requiring treatment, active cancer (excluding certain skin cancers), cardiovascular disease, congenital abnormalities, renal failure requiring dialysis, chronic respiratory disease, severe autoimmune disease requiring biologic therapy, insulin-dependent diabetes, immunodeficiency (including HIV/AIDS), recent major surgery or trauma, or use of medications within the past six months known to alter immune response, such as high-dose corticosteroids.

### Sample collection

All 63 donors received high-dose quadrivalent inactivated influenza vaccine (Fluzone HD) approved for use in individuals ≥65 years during the fall influenza season of 2022–2023 at the University of Connecticut Health at Farmington, CT. Blood samples were collected at six study visits, nasal swabs at two visits, and stool samples at two time points. Of 63 participants, we further studied 60 donors (24 men and 36 women) (three were excluded due to incomplete HAI titer data across required timepoints).

### Blood Processing and Sample Storage

Peripheral blood samples were processed within one hour of collection in BD Vacutainer tubes containing Acid Citrate Dextrose (BD). Peripheral blood mononuclear cells (PBMCs) were isolated using density gradient centrifugation with Lymphoprep (StemCell Technologies). Isolated PBMCs were resuspended in a cryoprotective solution consisting of 90% fetal bovine serum (FBS) and 10% dimethyl sulfoxide (DMSO) and subsequently stored in liquid nitrogen for long-term preservation. Plasma samples were obtained by centrifugation of whole blood and were stored at −80 °C until further use.

### Determination of Cytomegalovirus (CMV) Serostatus

CMV serostatus was determined using a CMV IgG ELISA kit (Aviva Systems Biology) according to the manufacturer’s instructions. Plasma samples were thawed at room temperature, diluted, and analyzed as recommended. Results were calculated based on the controls provided in the assay kit.

### Hemagglutination Inhibition (HAI) Assay

The 2022–2023 Northern Hemisphere influenza vaccine composition included the following H1N1, H3N2, and influenza B strains: A/Victoria/2570/2019 (H1N1), A/Darwin/9/2021 (H3N2), B/Austria/1359417/2021, and B/Phuket/3073/2013. Reference virus stocks corresponding to these strains were generated in embryonated chicken eggs, quality controlled, titrated, and subsequently used for HAI assays to measure homologous antibody responses. Plasma samples from vaccinees were incubated overnight with three volumes (relative to plasma volume) of receptor-destroying enzyme (RDE; #10753-482) for 16–18 h at 37°C. The following day, three volumes of 2.5% sodium citrate solution were added, and samples were heat-inactivated at 56°C for 30 min. Final plasma dilutions were adjusted to 1:10 in PBS by adding three volumes of PBS relative to plasma volume. Each virus stock was diluted to a final concentration of 8 HA units/50 µL in FTA hemagglutination buffer (BD Biosciences, #211248). 50 µL of RDE-treated plasma was serially two-fold diluted. Subsequently, 25 µL of each plasma dilution was incubated with an equal volume of virus containing 8 HA units/50 µL for 30 min at room temperature. Then, 50 µL of turkey red blood cells (RBCs) (Lampire Biological), diluted to 0.5% in HA buffer, were added and incubated for 45 min at 4°C. Assays were performed in duplicate to ensure reproducibility. HAI titers were determined as the reciprocal of the highest plasma dilution that completely inhibited hemagglutination. Samples with titers below the limit of detection (1:10) were assigned a value of 10 for statistical analyses.

### Antibody Response Ranking

For each of the four vaccine strains, donors were individually classified based on FDA criteria: seroconversion was defined as a ≥4-fold increase in HAI titer at day 35 relative to baseline, and seroprotection (high baseline) was defined as a baseline HAI titer ≥40. Donors who neither seroconverted nor met the seroprotection threshold were classified as non-responders for that strain. Of the 60 donors, 4 had high baseline titers (HAI ≥40) for two or more strains and did not seroconvert; these individuals were excluded from downstream analyses.

To capture cumulative vaccine responsiveness across the remaining 56 donors and all four strains, donors were ranked using a previously published multivariate dense ranking framework^30^. For each strain, participants were assigned a dense rank according to their fold change in HAI titer (day 35/baseline). Individual strain ranks were summed to generate an overall score, and dense ranking was applied to these scores to determine each donor’s final rank. Individuals who seroconverted to two or more strains were classified as strong responders (SR), whereas those seroconverting to one or no strains were classified as weak responders (WR). To independently validate the ranking, baseline-adjusted fold changes were computed using the maxRBA function with scoreFun set to ’sum’ from the titer package in R^31^, and Spearman correlation was used to assess concordance between the two metrics. For downstream multi-omic analyses, 10 SR were selected among the highest-ranked responders and 10 WR among the lowest-ranked non-responders, while representing men and women in each group (**Table. S1d**).

### NULISAseq Assay

NULISAseq assays were performed on plasma samples from the top 10 SR and 10 WR at baseline, day 1, and day 7 at Alamar Biosciences as previously described^29^. Plasma samples stored at −80 °C were thawed on ice and centrifuged at 2,200 × g for 10 minutes to remove debris. A total of 25 µL of plasma per sample was plated into 96-well plates and analyzed using Alamar Biosciences’ Inflammation Panel 250, which targets predominantly inflammation- and immune response–related cytokines and chemokines. The NULISAseq workflow was performed on the ARGO™ HT platform. Immunocomplexes were formed using DNA-barcoded capture and detection antibodies. These immunocomplexes were captured and washed on paramagnetic oligo-dT beads, followed by release into a low-salt buffer. Released complexes were subsequently captured and washed on streptavidin beads. Proximal DNA ends on each immunocomplex were then ligated to generate a DNA reporter molecule containing both target-specific and sample-specific barcodes. DNA reporter molecules were pooled, PCR-amplified, purified, and sequenced on the AVITI platform (Element Biosciences). Sequencing data were processed using the NULISAseq algorithm (Alamar Biosciences). Sample- and target-specific barcodes were quantified, allowing up to two base mismatches or one indel plus one mismatch. Intraplate normalization was performed by dividing target counts for each well by the corresponding internal control counts. Interplate normalization was conducted using interplate control (IPC) normalization, in which counts were divided by the target-specific median of the three IPC wells on each plate. Data were rescaled, incremented by +1, and log₂-transformed to generate NULISA Protein Quantification (NPQ) values for downstream statistical analyses. Of the 20 samples, one failed quality control and was excluded; the remaining 19 samples were used for downstream analyses.

Differences in protein abundance between SR and WR at each timepoint were assessed using the Mann-Whitney U test. Longitudinal changes within each group were evaluated using the Wilcoxon signed-rank test. Spearman correlation was used to assess the association between NPQ values and vaccine responsiveness rank. A p-value of <0.05 was considered statistically significant for all comparisons.

### Predictive Modeling of Protein Biomarkers

To identify cytokine and chemokine concentrations (NPQ values) that most strongly discriminated SR from WR at each timepoint, analyses were restricted to proteins with statistically significant SR/WR differences. For each protein, Cliff’s delta score was calculated as an effect size measure to quantify the strength of separation between response groups. Proteins were selected sequentially: the protein with the highest Cliff’s delta score was retained first, followed by iterative selection of proteins that provided the greatest additional separation when combined with previously selected proteins, for up to five proteins total.

The discriminatory performance of each protein combination was evaluated by calculating the area under the receiver operating characteristic curve (AUC). For individual proteins, AUC was estimated using binomial logistic regression modeling SR/WR status as a function of protein concentration. For combinations of two or more proteins, binomial logistic regression with 4-fold cross-validation was applied, as implemented in the glmnet package in R. Given the small sample size, all 19 donors were used in both model training and testing to avoid overfitting.

### ELLA Microfluidic Immunoassay

The levels of four cytokines (IFNγ, IL-8, IL-10 and TNFα) and two chemokines (CCL2 and CXCL10) were measured in plasma samples from the top 10 SR and 10 WR at baseline, day 1, day 7, and day 35 using Simple Plex assays run on the ELLA microfluidic immunoassay platform (ProteinSimple, San Jose, CA). Plasma samples, diluted 1:1 in sample diluent, were added to the ELLA microfluidic cartridges (human SPCKE-06 cartridge for IL-8/CXCL8, CCL2/JE/MCP-1, IL-10, CXCL10/IP-10/CRG-2, IFN-gamma 3rd gen, TNFalpha 2nd gen) according to manufacturer’s instructions. The ELLA system reported the mean values of the built-in triplicate analysis for each analyte using the manufacturer-calibrated standard curve.

Differences in cytokine and chemokine levels between SR and WR at each timepoint were assessed using the Mann-Whitney U test. Longitudinal changes within each group were evaluated using the Wilcoxon signed-rank test. A p-value of <0.05 was considered statistically significant for all comparisons.

### Flow Cytometry

Three cell-surface staining panels were used to characterize major immune cell populations including T cells, NK cells and monocytes, CD4+ T cell subsets, and B cell subsets in PBMCs from 10 SR and 10 WR at baseline, day 1, day 7, and day 35. Fluorescence-labeled antibody cocktails for each panel were premixed in BD Horizon Brilliant Stain Buffer (BD Biosciences) 10 min before staining. Antibody cocktails were added to 100-μl aliquots containing 1 × 10⁶ PBMCs and samples were incubated for 30 min on ice. Stained samples were washed twice to remove unbound antibodies and then stained with live/dead fixable dye (Aqua, 1:1,000, Thermo-Fisher) for 15 min at room temperature.

For the analysis of major immune cell populations, cells were stained with fluorochrome-labeled antibodies targeting the following surface markers: CD3 PerCP-Cy5.5 (clone UCHT1, BD Biosciences, 1:200), CD4 BV786 (clone SK3, BD Biosciences, 1:100), CD19 APC (clone HIB19, BD Biosciences, 1:20), CD14 AF700 (clone MSE2, BD Biosciences, 1:50), CD56 APC-Cy7 (clone HCD56, BioLegend, 1:100), CD16 BV570 (clone 3G8, BioLegend, 1:50), CD27 BV605 (clone O323, BioLegend, 1:50), CD8 PB (clone RPA-T8, BD Biosciences, 1:100), CD45RA ECD (clone 3H4, Beckman Coulter, 1:100), CD197 (CCR7) BV711 (clone G043H7, BioLegend, 1:20), IgD FITC (clone IA6-2, BD Biosciences, 1:25), and HLA-DR PE (clone L243, BioLegend, 1:100).

For the analysis of CD4+ T cell subsets, cells were stained with fluorochrome-labeled antibodies targeting the following surface markers: CD3 AF700 (clone UCHT1, BioLegend, 1:200), CD4 APC-Cy7 (clone OKT4, BioLegend, 1:100), CD183 (CXCR3) BV421 (clone GO25H7, BioLegend, 1:100), CD196 (CCR6) PE (clone 11A9, BD Biosciences, 1:100), CD185 (CXCR5) APC (clone J252D4, BioLegend, 1:100), CD279 (PD-1) FITC (clone MIH4, BD Biosciences, 1:100), CD278 (ICOS) PE-Cy7 (clone C398.4A, BioLegend, 1:200), and CD45RA ECD (clone 3H4, Beckman Coulter, 1:100).

For the analysis of B cell subsets, cells were stained with fluorochrome-labeled antibodies targeting the following surface markers: CD19 APC (clone HIB19, BD Biosciences, 1:20), IgD APC-H7 (clone IA6-2, BD Biosciences, 1:25), CD27 PE (clone M-T271, BD Biosciences, 1:50), CD20 PO (clone HI47, ThermoFisher, 1:25), CD38 AF700 (clone HIT2, BioLegend, 1:50), CD138 PerCP-Cy5.5 (clone MI15, BD Biosciences, 1:20), CD185 (CXCR5) APC (clone RF8B2, BD Biosciences, 1:100), CD21 PE-Cy7 (clone B-ly4, BD Biosciences, 1:50), CD24 BV711 (clone ML5, BioLegend, 1:50), and CD11c V450 (clone B-ly6, BD Biosciences, 1:50). Full antibody details are provided in **Table S6**. Cells were acquired on an LSR Fortessa X-20 (BD Biosciences) and data were analyzed using FlowJo software (BD Biosciences).

For sorting of CD4⁺ T cell and CD19⁺ B cell subsets, PBMCs isolated from blood samples of influenza vaccinees collected at day 7 post-immunization were stained with the following antibodies: anti-CD3 (clone UCHT1, BD Biosciences; 1:200), anti-CD4 (clone RPA-T4, BD Biosciences; 1:100), anti-CD19 APC (clone HIB19, BD Biosciences; 1:20), anti-CD45RA ECD (clone 3H4, Beckman Coulter; 1:100), anti-CXCR5 (clone RF8B2, BD Biosciences; 1:100), anti-CXCR3 (clone G025H7, BioLegend; 1:100), anti-CD27 BV605 (clone O323, BioLegend; 1:50), anti-IgD APC-H7 (clone IA6-2, BD Biosciences; 1:25), and anti-PD-1 (CD279) BV786 (clone EH12.1, BD Biosciences; 1:100). Tfh (CD3⁺CD4⁺CD45RA⁻CXCR5⁺), Th10 (CD3⁺CD4⁺CD45RA⁻CXCR5⁻CXCR3⁺PD-1^hi^), and memory B cells (CD3⁻CD19⁺IgD⁻CD27⁺) were sorted using a BD FACSAria Fusion cell sorter equipped with a 100-µm nozzle. Sorted cells were collected into cold RPMI 1640 medium supplemented with 10% fetal bovine serum. Sorting gates were defined as shown in **Fig. S10**. Post-sort purity was consistently greater than 98%.

Differences in cell subset frequencies between SR and WR at each timepoint were assessed using the Mann-Whitney U test. Longitudinal changes within each group were evaluated using the Wilcoxon signed-rank test. A p-value of <0.05 was considered statistically significant for all comparisons.

### T-B coculture assay

B cell helper function of Th10 cells was assessed using a T-B coculture assay. Th10 cells (CD3⁺CD4⁺CD45RA⁻CXCR5⁻CXCR3⁺PD-1^hi^ were sorted from PBMCs of influenza vaccinees at day 7 post-immunization. A total of 2 × 10⁴ Th10 cells per well were cocultured with 2 × 10⁴ autologous memory B cells (CD3⁻CD19⁺CD27⁺IgD⁻) per well for 6 days in RPMI 1640 medium (Gibco) supplemented with 1% L-glutamine (Sigma), 1% penicillin/streptomycin (Sigma), 1% sodium pyruvate (Sigma), 1% non-essential amino acids (Sigma), and 10% heat-inactivated fetal bovine serum (GemCell). Cocultures were performed in 96-well U-bottom plates in the presence of highly purified staphylococcal enterotoxin B (SEB; 0.5 µg/mL; Toxin Technology). T follicular helper (Tfh) (CD3⁺CD4⁺CD45RA⁻CXCR5⁺) cells were cocultured with memory B cells under the same conditions and used as a positive control.

For the analysis of B cell differentiation after 6 days of culture, recovered B cells were stained with the following antibodies: anti-CD19 APC (clone HIB19, BD Biosciences; 1:20), anti-CD27 BV605 (clone O323, BioLegend; 1:50), anti-CD38 AF700 (clone HIT2, BioLegend; 1:50), and anti-CD20 BV570 (clone 2H7, BioLegend; 1:50). Viability was assessed using Fixable Viability Dye Aqua (Thermo Fisher Scientific; 1:1000). Samples were acquired on an LSRFortessa X-20 flow cytometer (BD Biosciences), and data were analyzed using FlowJo software (BD Biosciences). Plasmablasts were defined as CD38^hi^CD20⁻ cells within the live CD19⁺ B cell population. Absolute plasmablast numbers per well were calculated based on the frequency of live plasmablasts determined by flow cytometry and the total number of viable cells recovered per well at the time of harvest. For the measurement of antigen-specific antibody production, supernatants from cocultures were collected and stored at −80 °C until analysis. Fluzone-specific IgG antibodies were quantified using an in-house ELISA, as described below.

### ELISA for detection of antigen-specific IgG

Flat-bottom 96-well MaxiSorp microtiter plates (ThermoFisher Scientific) were coated with 100 µL per well of Fluzone (1 µg/mL) diluted in carbonate coating buffer (pH 9.5; BioLegend) and incubated overnight at 4 °C.

Following coating, plates were blocked with ELISA assay diluent containing bovine serum albumin (BSA; BioLegend) for 1 hour. Plates were then incubated with 100 µL per well of T-B coculture supernatants at room temperature for 2 hours. After washing, plates were incubated for 1 hour at room temperature with horseradish peroxidase (HRP)-conjugated goat anti-human IgG (Bethyl) at a final dilution of 1:75,000 in ELISA assay diluent. Following extensive washing, tetramethylbenzidine (TMB) substrate (BD Biosciences) was added and incubated for up to 30 minutes at room temperature. The reaction was stopped with 2 N H₂SO₄, and absorbance at 450 nm was measured using a SpectraMax plate reader (Molecular Devices).

### 5’ single-cell library preparation and sequencing

After preparation of PBMC suspensions, cells were washed and suspended in PBS containing 0.04% BSA and immediately processed for single-cell library preparation. Cell viability and cell counts were assessed using a LUNA-FL automated cell counter with AO/PI staining. For each sample, 29,000 cells were loaded onto the 10x Genomics Chromium platform with an expected recovery of 20,000 single cells. Single-cell capture, barcoding, reverse transcription, and library preparation were performed on the 10x Genomics Chromium X platform using Chromium GEM-X Single Cell 5′ Kit v3 chemistry according to the manufacturer’s protocol (CG000733)^25^. From each individual sample, amplified cDNA was used to generate paired 5′ gene expression and T-cell receptor V(D)J libraries.

cDNA and final sequencing libraries were assessed for quality and quantity using TapeStation 4200, Qubit Fluorometer, and KAPA qPCR. Libraries were prepared using Library Construction Kit C and Dual Index Kit TT Set A. Libraries were sequenced on an Illumina NovaSeq X Plus instrument using paired-end sequencing with a 28-10-10-90 asymmetric read configuration. Sequencing was performed using NovaSeq X+ 10B 100-cycle and/or NovaSeq X+ 25B 300-cycle reagent kits. Across samples, Cell Ranger-reported mean reads per cell ranged from 28,045 to 68,451 for gene expression libraries, and mean used reads per cell ranged from 6,825 to 45,798 for V(D)J libraries.

### 5’ single-cell raw data processing

Illumina base call files were converted to FASTQ files, and paired gene expression and V(D)J FASTQ files were processed using the Cell Ranger multi pipeline v8.0.0. Gene expression reads were aligned to the 10x Genomics human GRCh38 reference, refdata-gex-GRCh38-2020-A, and TCR V(D)J reads were aligned to refdata-cellranger-vdj-GRCh38-alts-ensembl-5.0.0. Downstream analysis was performed using the filtered gene expression matrices and paired TCR contig annotations generated by Cell Ranger.

### 5’ scRNA-seq quality control, processing and annotation

Raw cellranger output was subjected to ambient RNA correction using SoupX^67^ (v1.6.2). Multiplets were identified and removed using Scrublet (v0.2.3)^68^ with an expected doublet rate of 6%, computed with parameters min_counts=2, min_cells=3, min_gene_variability_pctl=85 and n_prin_comps=30. Quality control was performed using Scanpy (v1.11.1), retaining cells expressing at least 200 genes and genes detected in at least 3 cells. Cells with mitochondrial transcripts comprising ≥20% of total counts were excluded. Batch correction across donors was performed using scvi-tools (version 1.3.0)^69^, and scVI-corrected representations were projected onto UMAP space and clusters were identified using the Leiden algorithm.

Cells were annotated through multiple rounds of iterative clustering and manual curation. In the first round, cells were partitioned into major immune lineages: myeloid cells, B cells, CD4+ T cells, and NK/CD8+ cytotoxic T cells, and further subclustered to resolve known PBMC subsets. The number of principal components and Leiden resolution were adjusted to optimize detection of known subsets. Clusters exhibiting markers from multiple lineages, red blood cell contamination, or elevated mitochondrial gene content were excluded. Remaining cells were re-clustered and annotated based on established immune cell marker genes. Within the plasmablast subset, immunoglobulin isotype identity was determined by assessing expression of immunoglobulin heavy chain constant region genes (*IGHM*, *IGHD*, *IGHG1*, *IGHG2*, *IGHG3*, *IGHG4*, *IGHA1* and *IGHA2*). Genes with log-normalised expression below 0.5 were excluded per cell; among the remaining genes, the isotype corresponding to the highest-expressed gene was assigned to that cell.

### Pre-processing and clustering of T cell subsets

T cells were sub-clustered from non-normalized count matrices, normalized to total counts per cell and log-transformed using Scanpy. Highly variable genes were identified, with TCR genes excluded prior to dimensionality reduction to prevent donor-specific TCR repertoire effects from driving clustering. Principal component analysis (n=100 components) was performed on highly variable genes, followed by Harmony based batch correction^70^. Neighbourhood graphs and UMAP embeddings were computed on Harmony-corrected representations, and clusters were identified using the Leiden algorithm. Cell clusters were annotated through iterative sub-clustering and manual curation using established marker genes, with the same QC exclusion criteria applied at each round. Within, CD4+ memory T cell subset, Th10 cells were identified by previously described transcriptional markers^53^, including *CXCR3, PDCD1, IL10, CD38, CXCR6, CCR2* and *CCR5*. Tph cells represent another population of helper cells that promote antibody production in autoimmune contexts. However, we could not identify a distinct cluster of Tph that expressed *CXCL13* within *PDCD1* expressing cells^71,53^.

Cell type frequencies were calculated as percentages within PBMCs and across broad cell lineages per donor. Longitudinal changes in cell frequency within SRs and WRs were assessed using a Wilcoxon signed-rank test. Between-group differences in cell frequency, or fold changes (relative to baseline), between SRs and WRs were assessed using a Mann-Whitney U test. Differentially expressed genes were identified from pseudobulked data using edgeR, with significance defined as a |log₂ fold change| > ±0.585 and FDR < 0.05.

### TCR (VDJ) processing and quality Control

Preprocessing of TCR data was performed using Scirpy pipeline (version 0.22.0)^72^. Data metrics were determined using functions ir.pp.index_chains and ir.tl.chain_qc functions with default parameters. Single pair chains were retained, excluding orphan chains, extra chains, and two full chains. Cell annotation labels were applied to TCR data from the gene expression and shared barcodes between the two modalities were retained for downstream analysis. Sequence distance metric were calculated using the ir.pp.ir_dist function using metric = ‘identity’ and sequence = ‘aa’. Clonotype clusters and clonotype network were determined with the criteria that both alpha and beta chain CDR3 amino acid sequences were identical, using the function ir.tl.define_clonotype_clusters and ir.tl.clonotype_network. Clonal expansion was calculated for each celltype at the sample level using the function ir.tl.alpha_diversity with metric = ‘gini_index’ parameter.

### Influenza-specific TCR annotation and quantification

TCR CDR3 amino acid sequence reference was compiled from multiple influenza-specific studies and large databases such as VDJDB and IEDB (**Table S7)**. Gamma/delta and ambiguous chains were excluded from reference as our TCR assay captured alpha and beta chains. From VDJDB, sequences associated with Homo Sapiens as host species and influenza A and B as the epitope species were retained. Further filtering was performed by retaining sequences that had a confidence score greater than 0. To predict antigen specificity in our cohort, we used a publicly available TCRMatch tool^73^ with a threshold score of 0.95. Specificity was determined if either the alpha or the beta chain was predicted to be influenza specific.

Clones were determined for each celltype using paired amino acid CDR3 sequence identity. Influenza-specific expanding clones were calculated as the log2 fold change in clonotype abundance relative to baseline after adding one pseudocount and clones were classified as expanding at day 7 if they showed a positive fold change and contained at least 2 cells. The frequency of influenza-specific expanding clonotypes at day 7 was then calculated for each donor relative to the total clonotype repertoire within the corresponding cell type.

### cTfh cells sorting

PBMCs were thawed and 1 million cells from each subject were aliquoted for cell hashing and individual subject demultiplexing. PBMCs were pooled together for CD4 T cell enrichment using EasySep Release Human CD4 Positive Selection Kit (Stem Cell). Enriched CD4 T cells were stained with 5uL per test of the following fluorescent antibodies: anti-human CD3 Antibody BV510 (Biolegend 317332), anti-human CD4 Antibody BV650 (Biolegend 317436), anti-human CD185 (CXCR5) Antibody BV711 (Biolegend 356934), and Spark PLUS UV™ 395 anti-human CD45RA (Biolegend 304190). Cells were also stained with 1uL per test of the following oligonucleotide tagged antibody for CITE-seq: anti-human/mouse/rat CD278 (ICOS) Antibody TotalseqC0171 (Biolegend 313553), anti-human CD183 (CXCR3) Antibody TotalseqC0140 (Biolegend 353747), anti-human CD196 (CCR6) Antibody TotalseqC0143 (Biolegend 353440), anti-human CD279 (PD-1) Antibody TotalseqC0088 (Biolegend 329963), anti-human HLA-DR Antibody TotalseqC0159 (Biolegend 307663), anti-human CD38 Antibody TotalseqC0410 (Biolegend 356637). Individual subject aliquots were stained with 5uL per test anti-human CD3 Antibody BV510 (Biolegend 317332), anti-human CD4 Antibody BV650 (Biolegend 317436), and 1uL per test unique TotalSeq-C anti-human hashtag (Biolegend). Pooled and inidivudally aliquoted cells were washed twice with FACS buffer (PBS supplemented with 2% FBS + 0.01% NaN_3_. Immediately before sorting, individual subject aliquots were pooled and together for sorting of CD3^+^ CD4^+^ cells for subject demultiplexing. Cells were sorted using Aurora CS spectral cell sorter (Cytek Biosciences) using SpectroFlo CS Version 1.4.0 (Cytek Biosciences). CD4 cTfh (CD3^+^ CD4^+^ CD45RA^-^ CXCR5^+^) from subjects enrolled in the trial were sorted from pre-vaccination and day 7-post vaccination samples.

### Library preparation and sequencing for CITE-seq

Sorted cTfh were partitioned into nanoliter-scale Gel Bead-In-Emulsions (GEMs) to achieve single cell resolution using the 10x Genomics Chromium Next GEM Single Cell 5’ HT Reagent kits v2 and Chromium Controller X according to the manufacturer’s instruction (CG000424 Rev B 10xGenomics). The sorted single cells were processed according to 5’ gene expression and T cell receptor (TCR) enrichment instructions (CG000424 Rev B 10xGenomics) to prepare the libraries for sequencing. Libraries were sequenced using an Illumina NovaSeq X Plus at Weill Cornell Medicine Genomics Core Facility. 5’ Gene expression libraries, TCR, and feature barcode were demultiplexed using cellranger mkfastq pipeline. FASTQ reads were mapped to the human genome (GRCh38-2020-A) either using a combination of cellranger count and cellranger vdj, or cellranger multi (v7.0.1 or newer). Donor identity of each cell was determined using an in-house hybrid demultiplexing^74^ approach that integrated results of cell hashing (hashtag) and a single nucleotide polymorphisms-based demultiplexing method (Souporcell)^75^ as previously described.

### CITE-seq quality control

Data generate from sorted cTfh cells were analyzed using Scanpy (version 1.10.4)^76^ and Muon (version 0.1.6) pipelines^77^. Cells that were classified as multiplets or unassigned to a specific sample were excluded from downstream analysis. Doublets were removed using Scrublet (version 0.2.3)^68^ with an expected doublet rate of 6%. Doublet scores were computed using scrub.scrub_doublets() with the following parameters: min_counts=2, min_cells=3, min_gene_variability_pctl=85, and n_prin_comps=30. Further quality control was performed by retaining cells that expressed at least 200 genes using sc.pp.filter_cells(min_genes=200) and filtering genes expressed in at least 3 cells using sc.pp.filter_genes(min_cells=3). Cells with mitochondrial transcripts comprising 20% or more of the total counts were excluded. The remaining high-quality cells were retained for downstream analysis.

### Batch correction, dimensionality reduction and clustering of CITE-seq

RNA modality was batch corrected using scvi-tools (version 1.3.0)^69^. scvi.Model.SCVI.setup_anndata function and the following parameters batch_key=’pool’, categorical_covariate_keys = [‘donor’,’sex’] and continuous_covariate_keys = [‘pct_counts_mt’,’pct_counts_ribo’,’age’] were used to create anndata object for this model. Variational Inference was calculated using scvi.model.SCVI with n_latent = 100. Counts were normalized to the total counts per cell using sc.pp.normalize_total() and log-transformed using sc.pp.log1p() functions. SCVI corrected PCs were used to compute neighborhood graph using sc.pp.neighbors function (n_pcs=60) and embedded using UMAP (sc.tl.umap(maxiter=50)). Clustering was performed using sc.tl.leiden() function.

ADT data was normalized using the using the CLR-normalization method from Muon function mu.prot.pp.clr(axis=1). Principal components were calculated using sc.tl.pca(svd_solver=’arpack’). Batch effects were corrected using Harmony integration through the Scanpy external API (sce.pp.harmony_integrate()), with max_iter_harmony=50. Harmony corrected PCs were used to compute neighborhood graph using sc.pp.neighbors function and embedded using UMAP (sc.tl.umap(maxiter=50)). Clustering was performed using sc.tl.leiden() function.

### Annotation and quantification of CITE-seq data

Depending on the granularity of celltype labels, RNA or ADT based processing and clustering were adapted along with mutpile rounds of clustering. cTfr cells were identified from RNA based clustering using well described markers, including *FOXP3*, *TIGIT*, *IL2RA*, and *IKZF2*. To analyze cTfh susbets in greater granularity, cTfh1, cTfh17 and HLA-DR+ cTfh cells were identified by ADT-based clustering using the cell-surface CXCR3, CCR6, and HLA-DR levels, respectively. These markers were quantified at the sample level by taking the mean of CLR normalized counts across cells.

### DOGMA-seq library preparation and sequencing

Cell suspensions from each sample were stained with TotalSeq-A oligo-conjugated antibody panels (BioLegend) targeting cell-surface proteins of interest and TotalSeq-A hashtag antibodies for sample multiplexing, according to the manufacturer’s protocol (BioLegend protocol CG000185: https://dx.doi.org/10.17504/protocols.io.8aahsae). A total of 80 samples were multiplexed into 20 pools, with four samples assigned to each pool using distinct hashtag labels. Stained cells were washed as pools and lysed under digitonin-based permissive conditions (“DIG”) as described^26^ to generate suspensions compatible with downstream single-nucleus multiome processing.

Cell quality and concentration were assessed on a LUNA FX7 automated cell counter. For samples with available pre-pool measurements, cell counts ranged from 0.33 × 10⁶ to 3.82 × 10⁶ cells per sample, and viability ranged from 51.2% to 94.8%. Transposition was performed for each multiplexed pool using the Chromium Next GEM Single Cell Multiome ATAC + Gene Expression Reagent Kit (10x Genomics, PN-1000283) according to the manufacturer’s protocol (#CG000338), modified per Mimitou et al^26^. to include a bridge oligo in the barcoding reaction enabling capture of ADT and HTO molecules into the ATAC fraction. Each multiplexed pool was processed as two paired Multiome RNA/ATAC library reactions with corresponding HTO and cell-surface protein libraries, generating 40 paired ARC library outputs across the 20 pools. Transposed cells were loaded onto Chromium Next GEM Chip J (10x Genomics, PN-1000234). Across completed ARC libraries, Cell Ranger ARC estimated 4,982–20,000 recovered cells per library pair, with a median of 8,493 cells. Single-cell capture, barcoding, and library preparation were performed on a 10x Genomics Chromium platform^25^.

Following GEM recovery, the ADT and HTO fraction was separated from the ATAC pre-amplification product via SPRI size selection and used as input for ADT and HTO library construction with indexed PCR primers as described in Mimitou et al., 2021, while gene expression and ATAC libraries were completed according to the standard Multiome protocol. All libraries were checked for quality by TapeStation 4200 (Agilent) and Qubit Fluorometer (Thermo Fisher), quantified by KAPA qPCR, and sequenced on an Illumina NovaSeq X+ 10B 100-cycle flow cell. Gene expression libraries were sequenced with a 28-10-10-90 asymmetric read configuration, and ATAC libraries were sequenced with a 50-8-24-50 read configuration. Across the 40 paired ARC libraries, gene expression libraries were sequenced to 39,542–166,142 mean raw reads per cell, with a median of 98,351 reads per cell, and ATAC libraries were sequenced to 42,698–175,567 mean raw read pairs per cell, with a median of 103,827 read pairs per cell. ADT and HTO libraries were sequenced with a 28-10-10-90 read configuration.

Illumina base call files for all libraries were converted to FASTQs using bcl2fastq v2.20.0.422 (Illumina). Paired gene expression and ATAC FASTQs were processed against the GRCh38 Cell Ranger ARC reference using the Cell Ranger ARC count pipeline v2.0.1 (10x Genomics). ADT and HTO FASTQs were processed against a custom feature reference using CITE-seq-Count v1.4.5, and resulting count matrices were merged with the gene expression and ATAC outputs by cell barcode for downstream analysis. Downstream analyses were performed on the merged gene expression, chromatin accessibility, ADT, and HTO count matrices.

### DOGMA-seq Data Processing and Analysis

Cellranger outputs were aggregated using cellranger-arc aggr (version 2.0.2). To remove ambient RNA, SoupX^67^ was run on gene expression modality. Doublets/multiplets in gene expression modality were predicted using Scrublet (version 0.2.3) with a threshold of 0.18 and Amulet (version 1.1) for ATAC modality^68,78^. Cells were retained for downstream analysis only if they were classified as singlets for both methods. Demultiplexing was performed using the R package deMULTIplex2^79^.

scRNA-seq and ADT data were processed using Muon and Scanpy^76,77^. ADT data were normalized using the dsb package and used in the annotation of cell subsets. Clustering was performed using scRNA-seq data after selecting highly variable genes using the Scanpy pipeline. Batch correction was performed using Harmony with “donor” as the batch key^70^. scATAC-seq data were processed and quality control (QC) metrics were assessed using SnapATAC2 from 10x fragment files^80^. Cell multiplets were identified using AMULET with a false discovery rate (FDR) threshold of <5% and excluded from downstream analyses^78^. Cells with fewer than 1,000 fragments or a transcription start site enrichment (TSSe) score below 5 were removed. After quality control, two samples from each time point were excluded. Cell subset labels were transferred from scRNA-seq to scATAC-seq data based on the intersection of cells across both modalities. Peak calling was performed on BAM files for each cell subset using MACS3 with the parameters --call-summits -q 0.05 --nomodel --extsize 200 --shift -100^81^. The peak matrix for each cell subset was constructed using SnapATAC2 with a paired-insertion counting strategy^82^.

Peaks were retained if they were detected in at least 2% of cells in any of the vaccine visit groups. The retained peaks were normalized using SNAIL (Smooth-quantile Normalization Adaptation for the Inference of co-expression Links)^83^. An epigenetic interferon score was derived by aggregating chromatin accessibility across ISG-associated peaks per donor. Transcription factor (TF) motif accessibility deviations were computed for each cell subset using ChromVAR with HOCOMOCO motifs ^38,84^. TF binding site occupancy was predicted using the TOBIAS pipeline (v0.14.0) with HOCOMOCO motifs^85^. Volcano plots of ChromVAR motif deviation scores calculated at the donor level, where deviation scores were averaged across cells within each donor. Statistical significance was assessed using an unpaired t-test performed at the single-cell level.

### Gene Set Enrichment Analysis

Gene set enrichment analysis (GSEA) was performed using the fgsea package in R^86^. Gene sets were obtained from MSigDB^87^ (version 7.5.1) and included the Hallmark collection, BIOCARTA, and KEGG pathway gene sets. For each cell subset, genes were ranked using a signed score computed as −log(p-value) × sign(log fold change), combining both the statistical significance and direction of differential expression. Pre-ranked GSEA was performed using fgsea with eps = 0 to enable precise p-value estimation. Pathways with an adjusted p-value < 0.005 and a normalized enrichment score (NES) > 1.5 were considered statistically significant.

### Independent Older Adult Cohort

To validate the cDC2 findings, an independent cohort of community-dwelling older adults (>65 years) from a previously published study was reanalyzed^6^. Participants received either standard-dose (SD, n=23) or high-dose (HD, n=28) quadrivalent inactivated influenza vaccine (QIV) during the 2021–2022 influenza season. Our multivariate dense ranking framework was applied to classify donors as SR or WR. In the HD group, all donors were classified as SR except for one individual, who was excluded from comparative analyses. cDC2 frequencies at day 1 post-vaccination were compared across SD-SR, SD-WR, and HD-SR groups using the Mann-Whitney U test. Spearman correlation was used to assess the association between cDC2 fold change at day 1 and responsiveness rank among HD-SRs.

### Independent Young adult cohort

To compare cDC2 transcriptional responses between younger and older adults, single-cell RNA-seq data from an independent cohort of 7 healthy younger adults (age range 22–39 years) who received standard-dose influenza vaccine (Fluzone SD, 2016-2017, 2017-2018) were preprocessed using the pipeline described above, with the exception of Harmony-based batch correction. DC subsets were identified by subclustering. For each donor, a transcriptional response score was computed at day 1 as the mean log-normalized expression of genes significantly upregulated in cDC2s of SR at day 1 (relative to baseline) in the primary cohort. Differences in scores between younger adults, SR and WR were assessed using a Mann-Whitney U test.

### Sound life cohort

Memory CD4+ T and B cell raw counts were obtained from the original publication^55^. These data provided by the authors had undergone quality control steps including doublet removal and filtering cells based on mitochondrial transcripts. We performed further quality control using Scanpy by retaining cells that express at least 200 genes and filtering genes that were expressed in less than 3 cells. Counts were normalized to the total read counts per cell using the sc.pp.normalize_total() function. Log-transformation was applied using sc.pp.log1p(). Highly variable genes were calculated using default settings using sc.pp.highly_variable_genes(). For B cells, genes associated with Ig isotype were removed from highly variable gene list to avoid isotype specific clusters. Principal components were calculated on highly variable genes with svd_solver = ‘arpack’ parameter. Batch correction was performed on ‘batch_id’ using sce.pp.harmony_integrate() function and max_iter_harmony=50. Harmony corrected PCs were used to compute neighborhood graph using sc.pp.neighbors(n_pcs=80) function and embedded using UMAP (sc.tl.umap(maxiter=50)). Clustering was performed using sc.tl.leiden() function.

Th10 cells were identified within memory CD4 T cell population by multiple rounds of clustering using previously described markers, including *CXCR3, PDCD1, IL10, CD38, CXCR6, CCR2* and *CCR5.* Plasmablasts were identified based on expression of *JCHAIN*, *PRDM1*, and *MZB1*.

## DATA AVAILABILITY

Raw genomics data (scRNA-seq, DOGMA-seq, CITE-seq) will be deposited in dbGaP under accession phs004793 upon completion of curator review and approval. Raw genomics data (scRNA-seq) for baseline samples are available in dbGaP under accession phs004450.v1.p1.

## CODE AVAILABILITY

Code is available on github: https://github.com/UcarLab/FluzoneHD

## Author contributions

P.W., A.G.-S., G.A.K., J.B. and D.U. designed the study and secured funding. G.A.K. and D.U. supervised the study. A.S.M. and S.R. contributed equally to this work as co-first authors and led the computational and multi-omic data analyses. Y.Y.Y. analyzed DOGMA-seq data. R.M. and K.K. processed blood samples and generated flow cytometry data. R.M. conducted co-culture experiments. H.J. contributed to flow cytometry data analysis. S.N. and P.W. generated CITE-seq data and plasmablast flow cytometry data. T.A. and A.R.-F. performed HAI titer assays. T.A. and A.G.-S. supervised antibody quantification analyses. L.K.P. coordinated participant recruitment, scheduling, and sample collection. D.K. contributed to cytokine data analysis. D.N.-B., V.P., P.W., A.G.-S., P.S., and J.B. contributed to data interpretation. A.S.M., S.R., J.B. and D.U. wrote the manuscript. All authors reviewed and revised the manuscript.

## Data Availability

All data produced in the present study are available upon reasonable request to the authors

## Acknowledgements

We thank JAX Single Cell core for their help with generating the single cell sequencing data. We thank Olivia Bart for help with dbGAP data upload. We thank members of the Ucar lab for critical feedback during the progress of the study. We thank the research participants who contributed to our work. ChatGPT (OpenAI; GPT-5–series models) and Claude (Anthropic; Opus 4.6–4.7) were used for revising and improving the clarity of language for this manuscript.

