## Supplementary Figure Legends for "Post-vaccination expansion of extrafollicular Th10 and regulatory Tfr cells distinguishes strong from weak influenza vaccine responses in older adults"

**ABSTRACT**

Despite the superior efficacy of high-dose influenza vaccines, over one-third of older adults fail to respond. Yet, the mechanisms underlying this impaired vaccine responsiveness remain poorly understood. Here, we performed longitudinal profiling of older adults (n=60) receiving high-dose influenza vaccination to identify immune programs associated with vaccine responsiveness. Strong responders exhibited a primed baseline immune state characterized by elevated plasma cytokines and chemokines, followed by enhanced IFN-γ responses and coordinated transcriptional and epigenetic activation of cDC2 cells at day 1. By day 7, CD4⁺ T-cell trajectories diverged: strong responders preferentially expanded influenza-specific activated cTfh1 (*CXCR5^+^ CXCR3^+^ ICOS^+^ CD38^+^*) and influenza-specific Th10 (*CXCR5⁻ CXCR3^+^ PD1^+^ IL10⁺*) cells, whereas weak responders expanded regulatory cTfr (*CXCR5⁺ FOXP3⁺*) cells. Th10 expansion correlated with plasmablast and antibody responses and was independently validated in a larger influenza vaccination cohort, including younger adults. Functionally, Th10 cells promoted memory B-cell differentiation into plasmablasts and production of influenza-specific IgGs. TCR analyses revealed minimal clonal overlap between Th10 and cTfh1 cells. Together, these findings identify divergent helper and regulatory CD4⁺ T cell programs associated with vaccine responsiveness and establish Th10 cells as a previously unrecognized component of vaccine-induced humoral immunity.

**Supplementary Figure Legends**

**Figure S1. Heterogeneous antibody responses to Fluzone HD in older adults.**

**a,** HAI titers at Pre, days 7 and 35 across all 60 donors for each of the four vaccine strains (A/H1N1, A/H3N2, B/Austria, B/Phuket). **b,** Stacked bar plots showing the proportion of donors classified as seroconverted (day 35/Pre ≥ 4), seroprotected (baseline HAI ≥ 40), or non-responsive (day 35/Pre < 4 and baseline HAI < 40) for each vaccine strain. **c,** Spearman correlations between antibody response rank and age, BMI, anti-CMV IgG titer, and Rockwood Frailty Index. **d,** Antibody response rank compared between females and males. **e,** HAI titers for the B/Phuket strain at Pre, days 7 and 35 in WR and SR (n = 10 each). Box plots display the median and IQR (25th–75th percentile); whiskers extend to ±1.5 × IQR. Exact p-values are shown; ns, not significant. Wilcoxon signed-rank test (two-sided; a); Mann–Whitney U test (two-sided; d, e); Spearman correlation (c).

**Figure S2. SRs exhibit a ‘primed’ baseline plasma proteome and enhanced IFN-γ responses.**

**a,** Box plots of baseline NPQ (normalized protein quantity) values for 25 plasma proteins significantly elevated in SR (n = 9) versus WR (n = 10). **b,** Spearman correlations between baseline NPQ values and antibody response rank for the 13 proteins most significantly correlated with antibody response rank. **c,** IFN-γ and TNF-α protein abundance (pg/mL) measured by ELLA microfluidic immunoassay at Pre, days 1, 7, and 35 in WR (n = 10) and SR (n = 10). Box plots display the median and IQR (25th–75th percentile); whiskers extend to ±1.5 × IQR. Exact p-values are shown; ns, not significant. Mann–Whitney U test (two-sided; a, c); Wilcoxon signed-rank test (two-sided) for longitudinal within-group comparisons (c); Spearman correlation (b).

**Figure S3. Strong interferon responses of SR in cDC2 at day 1.**

**a,** UMAP of all PBMCs from scRNA-seq (n = 956,539 cells) identifying ten major cell subsets: monocytes, DC & pDC, NK cells, CD4⁺ T cells, CD8⁺ T cells, naïve B cells, memory B cells, plasmablasts, and HSPCs. **b,** Dot plot showing expression of canonical marker genes of major cell subset; dot size represents the fraction of cells expressing the gene and color intensity represents mean expression. **c,** Longitudinal frequencies of total DC and monoDC in WR (n = 10) and SR (n = 10). **d,** Spearman correlations between day 1/Pre fold change of total DC, cDC2, and monoDC frequencies and antibody response rank. **e,** Spearman correlation between day 1/Pre fold change in cDC2 frequency and antibody response rank in an independent older adult cohort (Bonduelle et al., 2026; n = 28). **f,** Bar plot showing the number of DEGs at day 1 (relative to Pre) in cDC2, monoDC, and pDC subsets in SR. **g,** Heatmap of Z-score normalized expression of genes differentially expressed at day 1 (relative to Pre) in monoDCs of SR. Full list of differentially expressed genes is provided in **Table S4**. **h,** GSEA comparing SR versus WR at day 1 in cDC2, monoDC, and pDC subsets; bubble size reflects the number of leading-edge genes per gene set. **i,** Volcano plots of ChromVAR deviations in cDC2s at day 1/Pre in WR and SR; IRF family motifs are significantly enriched in SR. Box plots display the median and IQR (25th–75th percentile); whiskers extend to ±1.5 × IQR. Solid lines indicate the group mean at each timepoint. Exact p-values are shown; ns, not significant. Wilcoxon signed-rank test (two-sided; c); Mann–Whitney U test (two-sided; c).

**Figure S4. Day1 induction of transcriptional and epigenetic interferon responses in monocytes of SR.**

**a,** UMAP of monocytes from scRNA-seq (n = 248,547 cells) identifying three subsets: CD14⁺ monocytes, CD16⁺ monocytes, and intermediate monocytes. Feature plots show expression of canonical marker genes. **b,** Fold change (day 1/Pre) for total monocytes, CD14⁺, intermediate, and CD16⁺ monocyte subsets between WR (n = 10) and SR (n = 10). **c,** GSEA of top enriched pathways in SR versus WR at day 1; bubble size reflects the number of leading-edge genes. **d,** UMAP of monocytes from DOGMA-seq (n = 45,732 cells) identifying CD14⁺ and CD16⁺ monocyte subsets; feature plots show expression of CD14 and CD16. **e,** CD14⁺ monocyte interferon score (average read counts, log₂) at Pre, days 1, 7, and 35 in WR (n = 10) and SR (n = 10). **f,** Spearman correlation between transcriptional and epigenetic interferon scores in CD14⁺ monocytes. **g,** Volcano plot of ChromVAR deviations in CD14⁺ monocytes comparing SR versus WR at day 1. **h,** Heatmap of ChromVAR deviation scores for IRF and STAT family transcription factor motifs in CD14⁺ monocytes from WR and SR across Pre, days 1, 7, and 35; asterisks indicate significant enrichment in SR at day 1. **i,** Spearman correlation between IRF1 gene expression and IRF1 ChromVAR deviation in CD14⁺ monocytes at baseline and day 1 in WR and SR. **j,** Spearman correlation between ISG score (gene expression) and IRF1 ChromVAR deviation in CD14⁺ monocytes. Box plots display the median and IQR (25th–75th percentile); whiskers extend to ±1.5 × IQR. Solid lines indicate the group mean at each timepoint. Exact p-values are shown; ns, not significant. Mann–Whitney U test (two-sided; b, g); Wilcoxon signed-rank test (two-sided; e); Spearman correlation (f, i, j). n represents the number of biologically independent participants.

**Figure S5. Longitudinal B cell dynamics and early transcriptional activation in SR.**
**a,** Box plots of longitudinal naïve B cell, memory B cell, and plasmablast frequencies, shown as percentage of PBMCs, in WR (n = 10) and SR (n = 10) at Pre, day 1, and day 7. **b,** Bar plots showing the number of upregulated genes differentially expressed in SR at day 1 and day 7 relative to Pre in naïve B cells and memory B cells. **c,** Gene-set enrichment analysis of pathways significantly enriched in SR compared with WR at day 1 in naïve B cells and memory B cells. Dot size represents the number of genes contributing to each pathway. **d,** Left: UMAP of memory B cells (n = 16,195 cells) identifying unswitched memory B cells, switched memory B cells, DN2/atypical B cells, and activated B cells. Feature density plots show expression of selected marker genes and the day 7 transcriptional score of memory B cell DEGs. **e,f,** Box plots of longitudinal activated B cell and DN2 cell frequencies, shown as percentage of PBMCs, in WR (n = 10) and SR (n = 10) at Pre, day 1, and day 7. Box plots display the median and interquartile range (IQR; 25th–75th percentile), with whiskers extending to ±1.5 × IQR; individual data points are shown. Solid black lines indicate the group mean at each time point. Exact p-values are indicated on each panel. Wilcoxon signed-rank test (two-sided) was used for longitudinal within-group paired comparisons (a,e,f).

**Figure S6. SR show day 1 interferon signatures across T cell compartments.**

**a,** UMAP of T cells from scRNA-seq (n = 491,422 cells) identifying transcriptionally distinct T cell subsets. Feature density plots show expression of selected marker genes. **b,** Bar plots showing the number of upregulated and downregulated genes differentially expressed at day 1 relative to Pre in SR across major T cell compartments. **c,** Gene-set enrichment analysis of pathways enriched in SR compared with WR at day 1 across T cell compartments. Dot size represents the number of genes contributing to each pathway.

**Figure S7. Memory CD4 T cell subset annotation and vaccine-induced dynamics.**

**a,** UMAP of memory CD4⁺ T cells from scRNA-seq (n = 153,047 cells) identifying transcriptionally distinct memory CD4⁺ T cell subsets. Feature density plots show expression of selected marker genes used for subset annotation. **b,** Box plots of longitudinal memory CD4⁺ T cell subset frequencies in WR (n = 10) and SR (n = 10) at Pre, day 1, and day 7. **c,** Spearman correlation between day 7 activated cTfh1 expansion and antibody response rank. **d,** Spearman correlation between day 7 Th10 expansion and antibody response rank. **e,** Box plots of longitudinal CD4⁺CXCR5⁻CXCR3⁺PD-1⁺ Th10 cell frequencies quantified by flow cytometry in WR (n = 10) and SR (n = 10) at Pre, day 1, day 7, and day 35. **f,** Spearman correlation between day 7 activated cTfh1 expansion and day 7 Th10 expansion in WR. Solid black lines in box plots indicate the group mean at each time point. Box plots display the median and interquartile range (IQR; 25th–75th percentile), with whiskers extending to ±1.5 × IQR; individual data points are shown. Exact p-values are indicated on each panel; ns, not significant. Wilcoxon signed-rank test (two-sided) was used for longitudinal within-group paired comparisons (b and e); Spearman correlation was used for c, d and f.

**Figure S8. CITE-seq profiling of sorted cTfh cells.**

**a,** Experimental design and FACS gating strategy for sorting cTfh cells from WR (n = 10) and SR (n = 10) at Pre and day 7 for CITE-seq profiling. **b,** UMAP of sorted cTfh cells identifying transcriptionally distinct cTfh subclusters. Feature density plots show expression of selected marker genes used for subset annotation. **c,** UMAP of cTfh cells from CITE-seq (n = 97,152 cells), with violin plots showing cell-surface protein marker levels used to identify cTfh subsets. **d,** Box plots of longitudinal CD38 cell-surface protein levels in Tfh1 cells from WR (n = 10) and SR (n = 10) at Pre and day 7. Solid black lines in box plots indicate the group mean at each time point. Box plots display the median and interquartile range (IQR; 25th–75th percentile), with whiskers extending to ±1.5 × IQR; individual data points are shown. Exact p-values are indicated on each panel. Wilcoxon signed-rank test (two-sided) was used for longitudinal within-group paired comparisons (d).

**Figure S9. Th10 expansion is reproduced across age groups and vaccine formulations**

**a,** UMAP of memory CD4⁺ T cell subsets from the Sound Life cohort (n = 2,640,499 cells). Feature density plots show expression of selected marker genes used for subset annotation. **b,** Box plots comparing day 7 Th10 frequency fold change in young adults (Y; 25–35 years; n = 47) and middle-aged adults (M; 55–65 years; n = 45) after standard-dose influenza vaccination. **c,** UMAP of B cell subsets from the Sound Life cohort (n = 1,205,085 cells). Feature density plots show expression of selected B cell and plasmablast marker genes. **d,** Box plots of longitudinal plasmablast frequencies at day 0, day 7, and day 90 after standard-dose influenza vaccination. **e,** Box plots comparing day 7 plasmablast fold change across age groups. **f, g,** Spearman correlations between day 7 Th10 expansion and day 7 plasmablast expansion after Fluzone standard-dose vaccination or Fluarix standard-dose vaccination. Solid black lines in box plots indicate the group mean at each time point. Box plots display the median and interquartile range (IQR; 25th–75th percentile), with whiskers extending to ±1.5 × IQR; individual data points are shown. Exact p-values are indicated on each panel; ns, not significant. Wilcoxon signed-rank test (two-sided) was used for longitudinal paired comparisons (d) and fold-change comparisons relative to baseline where applicable (b, e); Spearman correlation was used for f and g.

**Figure S10. Sorting for co-culture studies.** Gating strategy of Th10 and cTfh and memory B cells to sort for co-culture experiments.
